# Sex-stratified genome-wide association meta-analysis of Major Depressive Disorder

**DOI:** 10.1101/2025.05.05.25326699

**Authors:** Jodi T Thomas, Jackson G Thorp, Floris Huider, Poppy Z Grimes, Rujia Wang, Pierre Youssef, Jonathan R.I. Coleman, Enda M. Byrne, Mark Adams, BIONIC consortium, The GLAD Study, Sarah E Medland, Ian B Hickie, Catherine M. Olsen, David C. Whiteman, Heather C. Whalley, Brenda WJH Penninx, Hanna M. van Loo, Eske M Derks, Thalia C. Eley, Gerome Breen, Dorret I Boomsma, Naomi R Wray, Nicholas G Martin, Brittany L Mitchell

## Abstract

There are striking sex differences in the prevalence and symptomology of Major Depressive Disorder (MDD). We conducted the largest sex-stratified genome wide association and genotype-by-sex interaction meta-analyses of MDD to date (Females: 130,471 cases, 159,521 controls. Males: 64,805 cases, 132,185 controls). We found 16 and eight independent genome-wide significant SNPs in females and males, respectively, including one novel variant on the X chromosome. MDD in females and males shows substantial genetic overlap with a large proportion of MDD variants displaying similar effect sizes across sexes. However, we also provide evidence for a higher burden of genetic risk in females which could be due to female-specific variants. Additionally, sex-specific pleiotropic effects may contribute to the higher prevalence of metabolic symptoms in females with MDD. These findings underscore the importance of considering sex-specific genetic architectures in the study of health conditions, including MDD, paving the way for more targeted treatment strategies.

## Introduction

Major Depressive Disorder (MDD) exhibits striking sex differences in both prevalence and clinical presentation. Globally, females are nearly twice as likely as males to experience MDD, with this disparity emerging around puberty and persisting into adulthood [1, 2]. This prevalence difference persists across a variety of forms of diagnosis, as well as across cultures and geographic borders [1, 3]. Beyond variation in prevalences, sex differences extend to symptomatology. Females tend to have a higher prevalence of atypical depression, characterized by symptoms such as weight gain, hypersomnia, and increased appetite, as well as immuno-metabolic depression, which is defined by immune-inflammatory pathophysiology and metabolic dysregulation and often overlaps symptomatically with atypical depression. In contrast, males with MDD more frequently exhibit anger, aggression, risk-taking behaviours, and substance use, with higher rates of comorbid substance use disorders [4, 5]. These differences suggest potential underlying biological and psychosocial mechanisms that contribute to MDD heterogeneity across sexes [6].

Multiple explanations have been proposed for this MDD heterogeneity across sexes, spanning behavioural, environmental, and biological domains. One potential explanation is variation in help-seeking and symptom reporting, as males are generally less likely to seek professional help or disclose symptoms, leading to under-diagnosis [7]. Environmental exposures also vary by sex; for example, females are more frequently exposed to sexual abuse and other forms of interpersonal violence, and experience structural forms of discrimination such as the gender wage gap, which may contribute to the higher prevalence of MDD in females [8, 9]. Additionally, biological mechanisms may underlie these differences, with research pointing to the roles of the immune system, neuroanatomy, neuroplasticity, stress and the hypothalamic-pituitary-adrenal (HPA) axis, and hormonal influences such as sex hormones and the hypothalamic-pituitary-gonadal (HPG) axis [10–14]. Together, these factors highlight the complexity of MDD heterogeneity across sexes and underscore the need for a multifaceted approach to understanding its underlying mechanisms.

Despite these numerous potential explanations, there has been limited replicated research clarifying the true aetiology of these sex differences in MDD. A key component of the biological mechanisms underlying these disparities could be differences in genetics. Research suggests that sex differences in human complex traits, including MDD, may arise from both sex-dependent and sex-specific genetic effects [15–18]. Sex-dependent effects imply that genetic variants could have differing effect sizes across sexes, with larger effects observed in one sex compared to the other. Additionally, sex-specific effects suggest that some of the genetic variants contributing to MDD may differ between males and females. Moreover, genetic variants on the X chromosome may play a crucial role in these sex differences, highlighting the need to consider sex chromosomes in genetic studies [19, 20].

There is mixed evidence for the role of genetics in the sex differences of MDD. A meta-analysis of five twin studies found no evidence for sex differences in the heritability of MDD [21]. However, other twin studies have shown higher heritability of MDD in females (40 – 51%) compared to males (29 – 41%), along with a genetic correlation between sexes that is significantly less than one [22–24]. More recently, genome-wide association studies (GWAS) have also provided mixed evidence for the existence of sex-dependent and sex-specific genetic effects contributing to these differences. For instance, a study using data from the UK Biobank reported a SNP-based genetic correlation between broad depression in males and females that was significantly less than one [17], however this was not the case in a large sex-stratified meta-analysis of MDD conducted by the Psychiatric Genomics Consortium [16]. In this same study, SNP-based heritability was estimated to be significantly higher in females than in males [16]. Furthermore, a cross-disorder (schizophrenia, bipolar disorder, MDD) genotype-by-sex interaction analysis identified a locus showing a significantly different association in females and males, further pointing to the role of sex-specific genetic factors [16]. These mixed results may stem from inconsistencies in cohort methodologies including case ascertainment [25]. Research has shown that genetic correlations can vary substantially across GWAS depending on the stringency and specificity of MDD phenotype definitions [26]. These findings underscore the need for consistent phenotyping approaches that align closely with diagnostic criteria to enhance the accuracy and comparability of genetic studies of MDD.

In this study we aimed to investigate whether sex differences in MDD can be explained, at least in part, by genetic effects. We conducted the largest sex-stratified GWAS and genome-wide genotype-by-sex interaction meta-analyses of MDD to date, using MDD cases primarily based on DSM (Diagnostic and Statistical Manual of Mental Disorders) criteria. Specifically, we examined whether genetic variants contribute to these differences through sex-dependent effects, sex-specific effects, or variants on the X chromosome. Additionally, we explored whether sex-specific pleiotropic effects between MDD and other phenotypes might help explain the observed differences in phenotypic presentation across sexes. These insights into sex-specific genetic mechanisms not only deepen our understanding of the aetiology of MDD but also emphasize the need for precision medicine strategies that account for sex differences.

## Results

### Sex-stratified GWAS

We conducted genome-wide association studies (GWAS) of Major Depressive Disorder (MDD) in five new cohorts and meta-analysed them with previously published GWAS meta-analysis summary statistics from Blokland *et al.* [16] for each sex separately. This resulted in a final sample size of 130,471 cases and 159,521 controls in females, and 64,805 cases and 132,185 controls in males (Supplementary Table 1). Our sex-stratified GWAS analyses identified 16 and eight independent genome-wide significant SNPs in females and males, respectively (Figure 1). In males, one novel SNP was identified on the X chromosome (rs5971319), which has not previously been associated with any depression phenotypes (Supplementary Tables 2 and 3).

**Figure 1.**
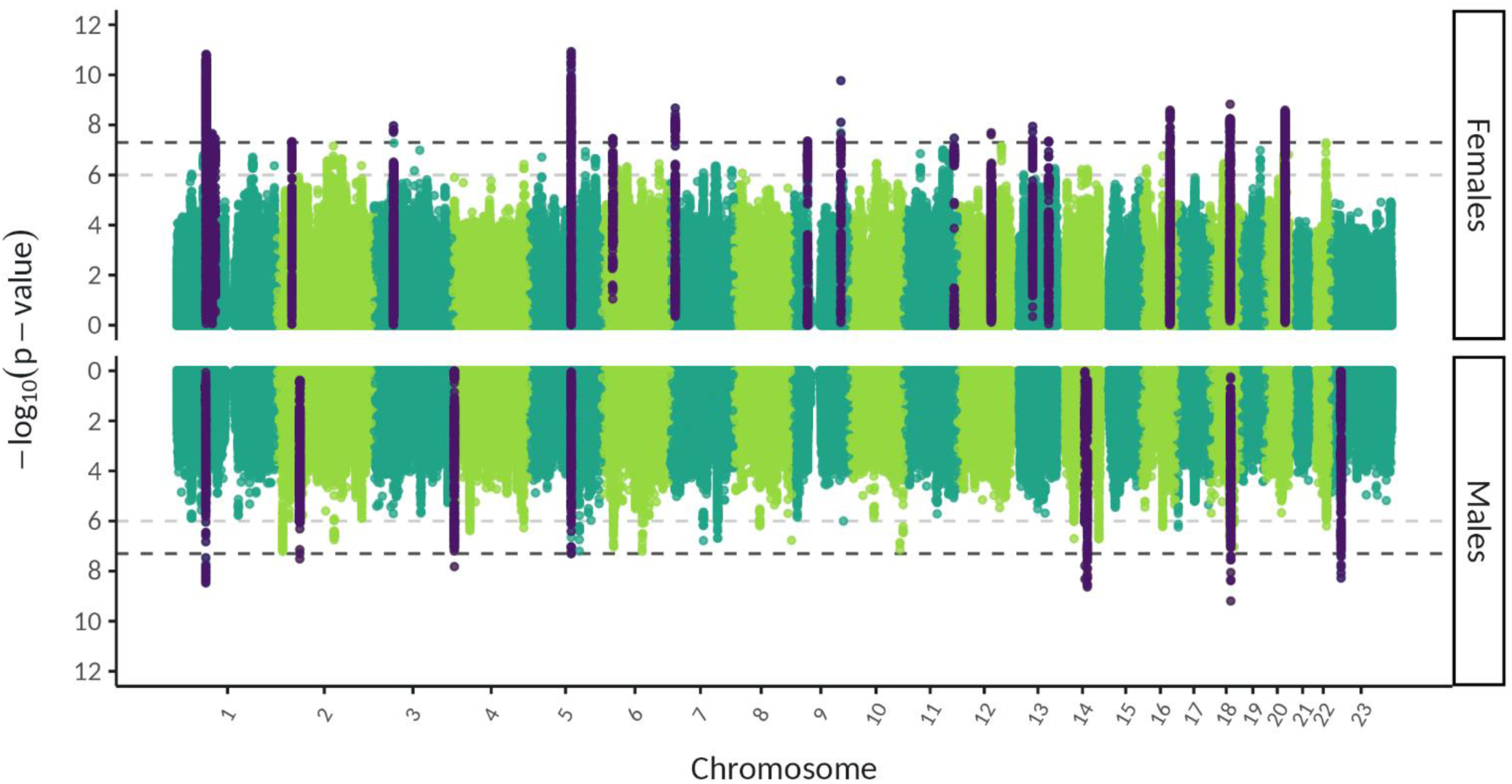
Miami plot of sex-stratified GWAS meta-analysis of MDD, with female and male meta-analyses shown on the top and bottom, respectively. The –log10 p values for each SNP are shown with positions according to human genome build 37 (GRCh37 assembly). Chromosome 23 is the X chromosome. The darker grey and lighter grey dotted horizontal lines indicate genome-wide significance (P = 5 x 10^-8^) and nominal significance (P = 1 x 10^-6^), respectively. SNPs in dark purple indicate the lead independent genome-wide significant SNPs, and any SNPs in linkage disequilibrium with them.

Our sex-stratified results showed high genetic correlation with the largest GWAS meta-analysis of MDD in both sexes combined [27] (Female-both sexes *r_g_* = 0.98 ± 0.01, Male-both sexes *r_g_* = 0.92 ± 0.02), as well as with two previously published sex-stratified GWAS of MDD [16, 17] (Female-Female *r_g_* = 0.91 – 0.96; Male-Male *r_g_* = 0.93 – 1.34) (Supplementary Figure 1, Supplementary Tables 4 and 5). Analysis of the genomic inflation factor (λ) and the Linkage Disequilibrium Score Regression (LDSC) intercept indicated little evidence for residual population stratification (Supplementary Figure 2). We tested the independent genome-wide significant SNPs (16 in females and eight in males) in our replication cohort, Generation Scotland. The number of SNPs with a concordant effect size direction was not significantly greater than expected by chance in both females (concordance = 69% [95% CI: 45 – 100%], H0: concordance = 0.5 [p = 0.11]) and males (concordance = 71% [95% CI: 34 –100%], H0: concordance = 0.5 [p = 0.23]) (Supplementary Tables 6 and 7). However, the replication cohort is small and the 95% confidence intervals are large suggesting the power of this replication is low.

#### Genetic Architecture of MDD in Females and Males

We investigated whether there was evidence for a sex difference in the genetic architecture of MDD by estimating sex-specific autosomal SNP-based heritability (*h*^2^_SNP_), polygenicity (π) and the selection parameter (*S*) using our sex-stratified meta-analysis results in SBayesS. *h*^2^_SNP_ was converted to the liability scale using a lifetime population risk of 0.2 and 0.1 in females and males, respectively. We found very strong evidence (100% posterior probability (PP)) that *h^2^*_SNP_ estimates are higher in females than males (*h*^2^_SNP_ female = 11.3% [95% highest posterior density interval (HPDI): 10.7 – 11.9%]; *h*^2^_SNP_ male = 9.2% [8.4 – 9.9%])) (Figure 2A). This suggests that the amount of variation in MDD risk explained by SNPs is higher in females. There was very strong evidence (100% PP) that MDD polygenicity is higher in females (π = 0.02 [0.015 – 0.024]) than males (π = 0.013 [0.009 – 0.017]), suggesting the number of SNPs contributing to MDD risk is higher in females (Figure 2B). We also applied univariate MiXeR [28] to quantify the polygenicity of MDD in females and males. The number of causal variants explaining 90% of MDD *h^2^_SNP_* was estimated to be 13,244 (SE = 1,120) for females and 7,111 (SE = 701) for males (Supplementary Table 8). These results also suggest MDD is more polygenic in females than males. Lastly, there was moderate evidence (79% PP) that the selection parameter was lower in males (*S* = –0.14 [-0.33 – 0.11]) than in females (*S* = –0.05 [-0.17 – 0.08]) (Figure 2C). This suggests that the negative relationship between effect size and minor allele frequency, an indicator of negative natural selection, could be stronger in males than females.

**Figure 2.**
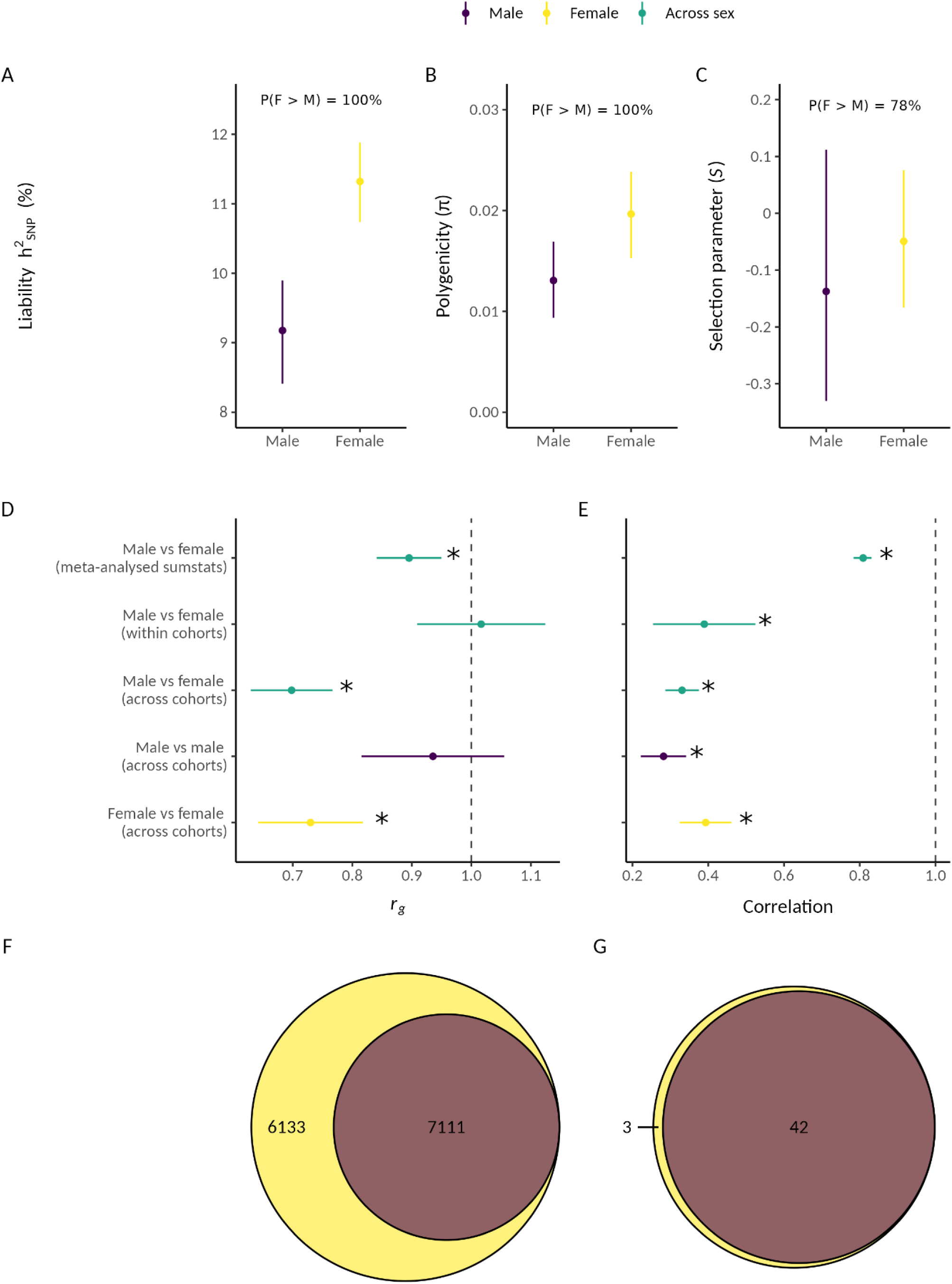
Comparisons of genetic architecture and polygenic overlap across the two sexes. A) Autosomal SNP-based heritability (*h^2^*_SNP_) on the liability scale using a population prevalence of 0.1 in males and 0.2 in females, B) Polygenicity, and C) Selection parameter using our sex-stratified meta-analysis results in SBayesS. D) Autosomal SNP-based genetic correlation (*r_g_*) between males and females using our sex-stratified meta-analysis results, and combination of all Markov chain Monte Carlo (MCMC) samples of *r_g_* estimated in all six male-female within cohort, 30 male-female across cohort, 15 male-male across cohort and 15 female-female across cohort combinations. E) Pearson correlation of the MDD effect sizes of SNPs known to be associated with sex-combined MDD for males vs females using our sex-stratified meta-analysis results, and meta-analysis of Pearson correlations estimated in all six male-female within cohort, 30 male-female across cohort, 15 male-male across cohort and 15 female-female across cohort combinations. F) Venn diagram depicting the number of causal variants explaining 90% of MDD *h^2^*_SNP_ in females only, males only, or both sexes, as identified by MiXeR. G) Venn diagram depicting the number of genomic regions that contain a causal variant for MDD in females only, males only or both sexes, as identified by gwas-pw. Females or female-female comparisons are in yellow, males or male-male comparisons in dark purple and female-male comparisons in green. For A) – C), as a Bayesian framework was used the points represent the mean posterior value, error bars are the 95% highest posterior density interval and percentages are the posterior probability that female value > male value. For D) and E), as frequentist statistics were used the points represent the mean while error bars are the 95% confidence interval and stars represent the *r_g_* / R being significantly different to 1 (after p-value correction for multiple tests).

We conducted a range of sensitivity analyses, including estimating *h^2^*_SNP_ using LDSC, exploring the impact of lifetime population prevalence and unscreened controls on *h^2^*_SNP_ (given evidence that MDD could be under-diagnosed in males [29]), and assessing the effect of differential power between the female and male GWAS and accounting for across-cohort heterogeneity [25] for *h^2^*_SNP_, π and *S*. Overall, our sensitivity analyses showed the same pattern of sex differences with a higher *h^2^*_SNP_ and polygenicity in females compared to males, however sex differences in the selection parameter did not remain in our sensitivity analyses. When the same population prevalence was used for males and females, we found that *h^2^*_SNP_ on the liability scale is similar across sexes, suggesting the sex difference in *h^2^*_SNP_ is driven by the prevalence difference (Supplementary Figure 3G). Refer to Supplementary Results A, Supplementary Figure 3 and Supplementary Tables 9 and 10 for more details.

#### Genetic Correlation of MDD Across Sexes

Autosomal SNP-based genetic correlation (*r_g_*) estimated from LDSC was used to determine whether there is evidence for sex-specific genetic effects, at the genome-wide level, contributing to MDD risk (Figure 2D). The *r_g_* between females and males was significantly less than one when using our sex-stratified meta-analysis results (*r_g_* = 0.90 ± 0.03, H0: *r_g_* = 1 [Z = –3.8, p_adj_(Benjamini-Hochberg) = 0.0003]). This result suggests that MDD variants are not fully shared across the sexes. To determine whether this *r_g_* < 1 reflects sex differences or general heterogeneity across cohorts, we also estimated *r_g_* between each pairwise combination of the two sexes by the six cohorts. Meta-analysis of the *r_g_* estimates from all 30 female-male across cohort comparisons was also significantly less than one (*r_g_* = 0.70 ± 0.03, H0: *r_g_* = 1 [Z = –8.6, p_adj_(B-H) = 3.7 x 10^-17^]). We then benchmarked these across-sex *r_g_* estimates against within-sex estimates; meta-analysis of the *r_g_* estimates from all 15 male-male across cohort comparisons was not significantly different from one (*r_g_* = 0.94 ± 0.06, H0: *r_g_* = 1 [Z = –1.06, p_adj_(B-H) = 0.36]), but the meta-analysis of all 15 female-female across cohort comparisons was significantly less than one (*r_g_* = 0.73 ± 0.04, H0: *r_g_* = 1 [Z = –6.01, p_adj_(B-H) = 4.56 x 10^-9^]). Furthermore, removing cohort variation by conducting a meta-analysis of the *r_g_* estimates from all six female-male within cohort comparisons showed a *r_g_* estimate not significantly different from one (*r_g_* = 1.02 ± 0.05, H0: *r_g_* = 1 [Z = 0.30, p_adj_(B-H) = 0.76]). Together, these results suggest that the female-male *r_g_* being significantly less than one could reflect heterogeneity between cohorts (especially among the female datasets) rather than true genome-wide sex differences.

We also investigated whether the MDD effect sizes (betas) are different across the sexes for SNPs known to be associated with MDD. The Pearson correlation was calculated between the standardised beta values of our male and female meta-analysis summary statistics, and between each pairwise combination of the two sexes by six cohorts, for the lead independent genome-wide significant SNPs from the largest GWAS meta-analysis of MDD [27] (Figure 2E, Supplementary Figures 4 – 6). The correlation between male and female MDD effect sizes was significantly less than one when using our sex-stratified meta-analysis results (R = 0.81 ± 0.02, H0: R = 1 [ Z = –9.34, p_adj_(B-H) = 1.23 x 10^-20^]) and for meta-analysis of the 30 male-female across cohort comparisons (R = 0.33 ± 0.02, H0: R = 1 [Z = –29.5, p_adj_(B-H) = 4.16 x 10^-191^]). Our benchmarking analyses were also all significantly less than one; meta-analysis of the 15 male-male (R = 0.28 ± 0.03, H0: R = 1 [Z = –23.6, p_adj_(B-H) = 9.91 x 10^-123^]) and 15 female-female (R = 0.39 ± 0.03, H0: R = 1 [Z = –17.35, p_adj_(B-H) = 3.17 x 10^-67^]) across cohort comparisons. Therefore, the female vs male correlations being significantly less than one could be due to the inherent heterogeneity of MDD rather than sex differences in the MDD effect sizes of SNPs associated with sex-combined MDD risk. However, removing cohort variation by conducting a meta-analysis of the correlations from all six female-male within cohort comparisons showed a correlation significantly less that one (R = 0.39 ± 0.07, H0: R = 1 [Z = –8.9, p_adj_(B-H) = 8.06 x 10^-19^]). This suggests that sex differences in genetic effects that are not fully explained by across cohort heterogeneity may exist within cohorts. A similar approach using the slope and intercept from linear regressions of male versus female effect size estimates also found similar conclusions (Supplementary Results B, Supplementary Figures 7 – 10).

#### Polygenic Overlap of MDD Across Sexes

Bivariate MiXeR [29] revealed that all 7,111 (SE = 701) causal variants for MDD in males were shared with MDD in females, with an additional 6,133 (SE = 988) variants unique to MDD in females and zero (SE = 0.0004) variants unique to MDD in males (Figure 2F). For those causal variants shared by females and males, the correlation of their effect sizes was 1.0 (SE = 6.4 x 10^-8^) (Supplementary Table 11). When accounting for the differential power across our sex-stratified GWAS we also found the same pattern (Supplementary Table 12, Supplementary Figure 3I). We also utilised gwas-pw [30] to identify autosomal regions of the genome containing MDD causal variants that are shared between females and males and regions that are sex-specific (Supplementary Figure 11). Gwas-pw identified 42 genomic regions that contain a causal variant for MDD shared by both sexes, three regions unique to females and zero regions unique to males, agreeing with the mixer results (Figure 2G). Within the 42 shared genomic regions, four possible causal risk variants were identified and mapped to multiple genes with two or more methods (*CYSTM1, PFDN1, HBEGF, SLC4A9, TLR4, CTD-2298J14.2, LRFN5, RAB27B*) (Supplementary Table 13). However, no possible causal risk variants were identified within the three female-specific genomic regions (Supplementary Table 14). Both our MiXeR and gwas-pw results suggest that the set of causal variants influencing MDD in males is a subset of those that influence MDD in females.

#### Functional annotation and analyses

We mapped each of the genome-wide significant SNPs from our sex-stratified GWAS meta-analysis to genes using positional, eQTL and chromatin interaction mapping (reporting genes with evidence from two or more mapping methods) (Supplementary Tables 15 and 16) and searched for previous SNP-phenotype associations in GWASCatalog. The female and male genome-wide significant SNPs mapped to different genes, with only one gene, *NEGR1*, being mapped to genome-wide significant SNPs from both sexes (Supplementary Table 17). The novel SNP identified in the male GWAS meta-analysis (rs5971319) has not previously been associated with any depression phenotypes but has previously been associated with educational attainment and vaginal microbiome relative abundance in the opposite effect size direction, and with neuroticism in the same effect size direction (Supplementary Tables 3 and 18). rs5971319 mapped to the gene *IL1RAPL1* (Supplementary Table 16).

We also conducted gene-based, gene-set and gene-property tests for our sex-stratified results using MAGMA v1.08 [31] within the SNP2GENE function in FUMA v1.5.2 [32]. 16 and 14 genes passed genome-wide significance (P < 2.53 x 10^-6^) from the female and male results, respectively. Only one gene, *DCC*, was significantly associated with MDD in both sexes (Supplementary Figure 12). In the female GWAS, SNPs were significantly enriched for genes involved in two biological processes; central nervous system neuron development (p-Bonferroni corrected = 0.036) and central nervous system neuron differentiation (p-Bonferroni corrected = 0.045). However, no gene sets were significantly enriched in the male GWAS. Gene-property analysis using 30 general tissue types (GTEx v8) identified SNPs from both the female and male stratified GWAS analyses as significantly enriched for gene expression in brain tissue, while pituitary tissue was significantly enriched only in the female SNPs (Supplementary Figure 13A). Furthermore, using 53 tissues types (GTEx v8), female and male SNPs were significantly enriched for gene expression in the cortex and frontal cortex, caudate, putamen and nucleus accumbens basal ganglia, hippocampus, amygdala, hypothalamus and anterior cingulate cortex. Female, but not male, SNPs were significantly enriched for gene expression in the cerebellum and cerebellar hemisphere (Supplementary Figure 13B). It is likely the differential power across the female and male GWAS meta-analysis results led to the difference in significant results for the gene-set and tissue analyses.

### Genome-wide genotype-by-sex interaction analysis

We tested for a significantly different association of SNPs with MDD across sexes by meta-analysing genome-wide genotype-by-sex interaction analyses (GxS) from each of the same five cohorts and the results from Blokland *et al.* [16]. No SNPs reached genome-wide significance, however four independent SNPs were nominally significant (P < 1 x 10^-6^) (Figure 3). The genomic inflation factor (λ) and the LDSC intercept indicated little evidence for residual population stratification (Supplementary Figure 14). The results for the X chromosome non-pseudoautosomal region were very similar when using full and no dosage compensation (Supplementary Figure 15). When restricting the GxS analysis to only the genome-wide significant SNPs found by the largest MDD GWAS meta-analysis [27] no SNPs reached significance for GxS (after Bonferroni multiple-testing correction) (Supplementary Figure 16). For more details about the nominally significant GxS SNPs refer to Supplementary Results C, Supplementary Figures 17 – 19, and Supplementary Tables 19 and 20.

**Figure 3.**
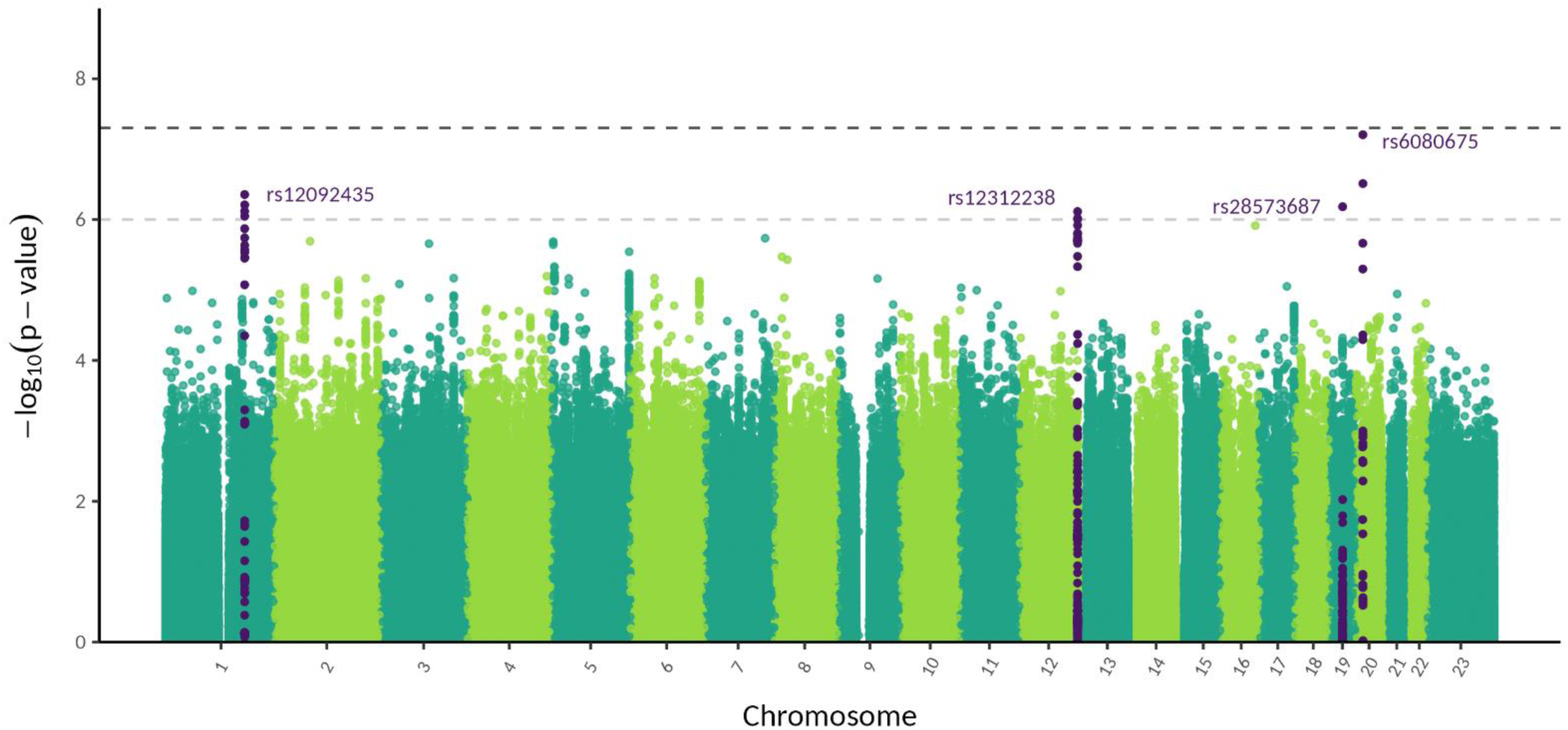
Manhattan plot of genome-wide genotype-by-sex interaction meta-analysis for MDD. The –log10 p-values for each SNP are shown with positions according to human genome build 37 (GRCh37 assembly). Chromsome 23 is the X chromosome. The darker grey and lighter grey dotted horizontal lines indicate genome-wide significance (P = 5 x 10^-8^) and nominal significance (P = 1 x 10^-6^), respectively. SNPs in dark purple indicate the lead independent nominally significant SNPs, and any SNPs in linkage disequilibrium with them.

### Sex-specific pleiotropic effects

#### Genetic Correlations

We used LDSC to identify genome-wide autosomal SNP-based genetic correlations (*r_g_*) between our sex-stratified MDD GWAS meta-analysis results and a range of other psychiatric disorders, metabolic traits and substance use traits (Figure 4A, Supplementary Table 21). Attention-deficit hyperactivity disorder (ADHD) was the only psychiatric condition with a sex-specific genetic correlation; there was a significantly higher genetic correlation between sex-combined ADHD and MDD in females compared to with MDD in males (Z = 3.34, p_adj_(Benjamini-Hochberg) = 0.003) (Supplementary Tables 22 and 23).

**Figure 4.**
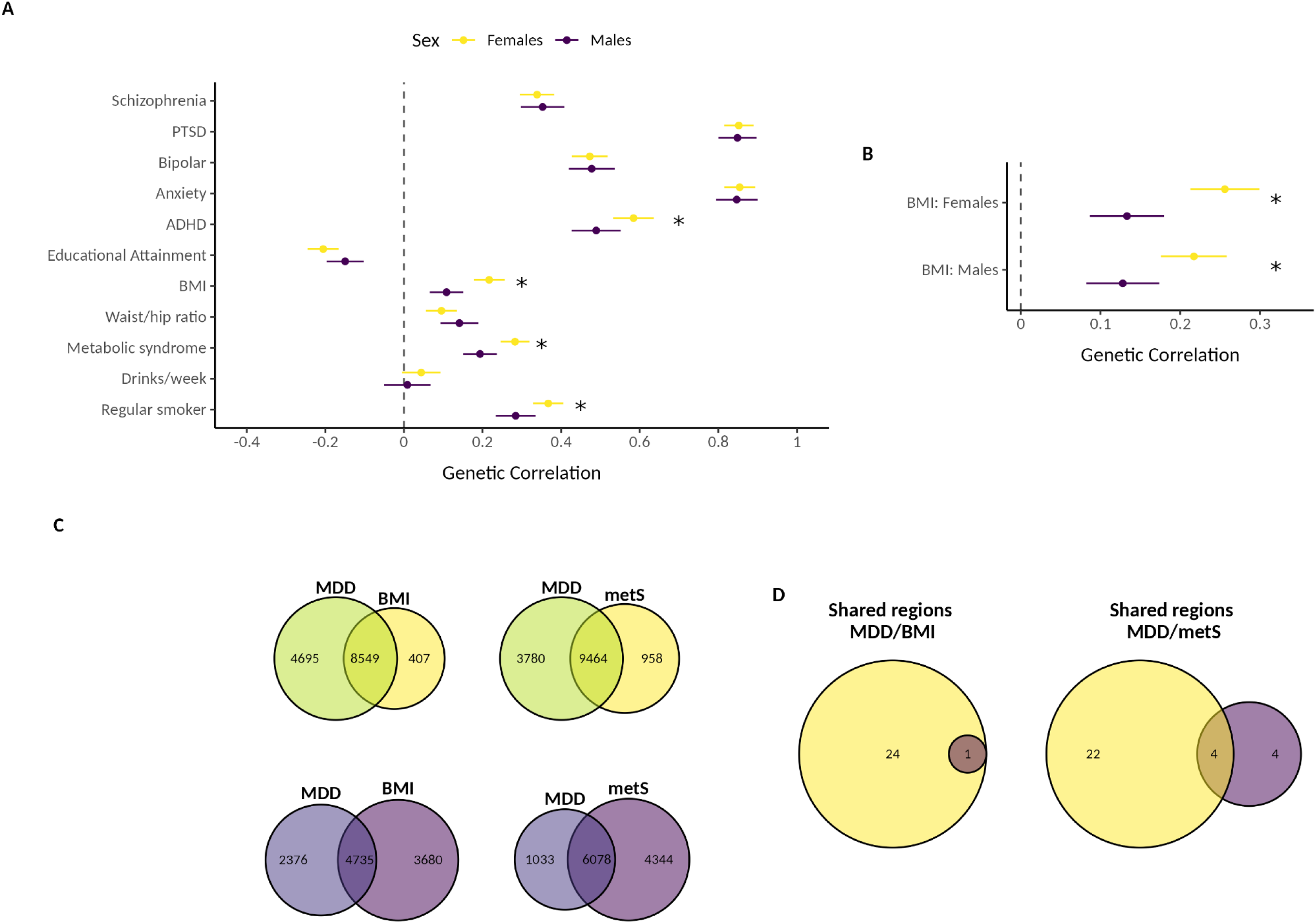
A) Genetic correlations of MDD in females and males with other psychiatric disorders, metabolic and substance use traits. B) Genetic correlations of MDD in females and males with BMI in females and males. C) Venn diagrams depicting the number of causal variants explaining 90% of *h^2^*_SNP_ in MDD only, BMI/metS only, or both traits as identified by MiXeR in females (yellows) and males (purples) D) Venn diagrams depicting the number of genomic regions that contain a causal variant for both MDD and BMI/metabolic syndrome in females only, both sexes, or males only as identified in gwas-pw. Females are in yellow and males in dark purple. * = significantly different genetic correlation between MDD and the other trait across sexes. PTSD = post-traumatic stress disorder, ADHD = attention deficit hyperactivity disorder, BMI = body mass index, Waist/hip ratio = waist to hip ratio adjusted for BMI, metS = metabolic syndrome.

There was also a significantly higher genetic correlation between sex-combined metabolic traits (body mass index (BMI) and metabolic syndrome) and MDD in females compared to with MDD in males (BMI: Z = 5.64, p_adj_(B-H) = 1.86 x 10^-7^, metabolic syndrome: Z = 4.35, p_adj_(B-H) = 1.38 x 10^-5^). As there is a well-powered sex-stratified GWAS of BMI, we also tested the genetic correlation of MDD in females and males with BMI in females and males (Supplementary Tables 24 and 25). Female MDD had a significantly higher genetic correlation with female BMI than male MDD had with female BMI (Z = 5.22, p_adj_(B-H) = 3.56 x 10^-7^). Female MDD also had a significantly higher genetic correlation with male BMI than male MDD had with male BMI (Z = 3.72, p_adj_(B-H) = 0.0002) (Figure 4B). Lastly, the genetic correlation of sex-combined regular smoking with MDD was significantly higher in females compared to males (Z = 2.85, p_adj_(B-H) = 0.01) (Figure 4A).

#### Polygenic Overlap

We used MiXeR to quantify the polygenic overlap between MDD in females/males and the metabolic traits BMI and metabolic syndrome (Figure 4C). The number of causal variants explaining 90% of *h^2^_SNP_* was estimated to be relatively similar for female and male BMI (female = 8,956 (SE = 232); male = 8,415 (SE = 237), while the number of causal variants estimated for metabolic syndrome (sex combined) was 10,422 (SE = 339) (Supplementary Table 26). Overall, female MDD had larger polygenic overlap with metabolic traits (female MDD-female BMI: n_shared_ = 8,549 ± 368; female MDD-sex combined MetS: n_shared_ = 9,463 ± 803) compared to male MDD (male MDD-male BMI: n_shared_ = 4,735 ± 689; male MDD-sex combined MetS: n_shared_ = 6,078 ± 969) (Supplementary Table 27).

We also utilised gwas-pw to identify sex-specific autosomal genomic regions containing causal risk variants shared between MDD and metabolic traits (Figure 4D). In both sexes, we identified one genomic region containing a causal variant for both MDD and BMI, which mapped to the genes *FTO* and *IRX3* (Supplementary Table 28). Of the 24 female-specific MDD/BMI pleiotropic regions identified, possible causal risk loci mapped to multiple genes (*DYNC1I2*, *HTT*, *MSANTD1*, *ANAPC4* and *DENND1A*) (Supplementary Table 29). No regions were identified as unique to MDD/BMI in males. We identified four genomic regions that contain a causal variant for both MDD and metabolic syndrome shared by both sexes, which also mapped to the *FTO* gene (Supplementary Table 30). Of the 22 female-specific MDD/metabolic syndrome pleiotropic regions, possible causal risk loci mapped to multiple genes (*GPC6, SHISA9, RP11-154H12.3, KCNG2, TXNL4A, RBFA* and *ADNP2*) (Supplementary Table 31). Lastly, from the four male-specific MDD/metabolic syndrome pleiotropic regions we identified one possible causal risk variant which mapped to the gene *ANKK1* (Supplementary Table 32).

## Discussion

Sex differences in the prevalence and symptomology of Major Depressive Disorder (MDD) are well-documented. Understanding these differences is crucial for uncovering underlying biological mechanisms and developing more targeted treatments [1, 4, 14, 33]. Here, we tested the following hypotheses: genetic variants may contribute to sex differences in MDD via 1) sex-dependent effects in which genetic variants have differing effect sizes for MDD across sexes, 2) sex-specific effects in which genetic variants contributing to MDD are different across sexes, and/or 3) the presence of genetic variants for MDD on the X chromosome. We also tested the hypothesis that sex-specific pleiotropic effects between MDD and other phenotypes may contribute to sex differences in the phenotypic presentation of MDD. To do this, we conducted the largest sex-stratified genome wide association and genotype-by-sex interaction meta-analyses for MDD to date.

In our GWAS of MDD in males we identified one novel independent genome-wide significant SNP (rs5971319). Notably, this novel SNP is located on the X chromosome and maps to the gene *IL1RAPL1* which is involved in the hippocampal memory system and is linked to intellectual disability [34]. A SNP in linkage disequilibrium (LD) with rs5971319 has previously been associated with educational attainment in the opposite direction. This is consistent with previous observations of the negative causal relationship between educational attainment and MDD risk [35]. The identification of this novel locus, despite having a substantially smaller sample size than the largest MDD GWAS meta-analysis (with females and males combined) [27] highlights the utility of sex-stratified analyses.

Our results provide evidence for sex differences in the heritability of MDD. Autosomal SNP-based heritability (*h^2^*_SNP_) was higher in females than males, and remained consistent in our sensitivity analyses accounting for the differential power across sexes and for across-cohort heterogeneity. This result reflects previous findings in a sex-stratified GWAS [16] and some twin studies [22–24], and suggests there may be a greater genetic contribution to MDD risk in females or alternatively a stronger environmental influence on MDD risk in males. We found that the sex difference in *h^2^*_SNP_ may be driven by the prevalence difference, as when the same population prevalence was used across sexes *h^2^*_SNP_ on the liability scale was similar in females and males. It is possible that MDD must be more severe in males compared with females for an individual to cross the liability threshold and for MDD to manifest. This increased male severity could explain the lower prevalence of MDD in males. As more severe phenotypes tend to have a higher heritability [36], it could also explain the disappearance of the sex difference when *h^2^*_SNP_ was calculated on the liability scale assuming equal population prevalence across sexes. Another explanation for the lower prevalence in males could be due to under-reporting and under-diagnosis of MDD in males [7, 37]. If this is the case, the true male population prevalence may be higher than reported and controls may include unidentified cases, resulting in an underestimation of male *h^2^*_SNP_ [38]. However, in our analyses *h^2^*_SNP_ remained higher in females even when accounting for up to 30% unscreened male controls, and a corresponding increase in male population prevalence, when compared to females with a population prevalence of 20% and no unscreened controls. Thus, *h^2^*_SNP_ could be higher in females even with under-reporting and under-diagnosis in males, although further research is needed to quantify this under-diagnosis.

The origin of the MDD prevalence difference across sexes could also affect interpretation of our heritability findings. If the higher prevalence of MDD in females is predominantly driven by environmental factors, such as trauma and structural forms of discrimination [8, 9], the sex difference in *h^2^*_SNP_ may arise from the statistical relationship between prevalence and liability scale *h^2^*_SNP_ rather than underlying sex differences in genetic architecture. However, if the prevalence difference is a consequence of genetic or biological factors, the difference in *h^2^*_SNP_ could indeed signal genuine sex differences in genetic architecture. Importantly, the presence of gene–environment correlations mean that differences in prevalence driven by environmental exposures may still partially reflect underlying genetic effects. Beyond heritability, we found evidence for a higher polygenicity for MDD in females compared to males. Our polygenic overlap analyses also showed that MDD causal variants in males are a subset of causal variants in females. These results remained consistent in our sensitivity analyses accounting for the differential power across sexes. This suggests that the higher *h^2^*_SNP_ in females may reflect, at least in part, distinct genetic architectures across sexes. Overall, our results suggest sex differences in the genetic architecture of MDD with a greater genetic burden for MDD risk in females, which may be driven by a higher number of MDD causal variants and the presence of female-specific SNPs, supporting the sex-specific hypothesis.

We found substantial overlap in genetic variants associated with MDD between male and female MDD. Genetic variants that influence MDD in both sexes had a near-perfect positive correlation. On the other hand, we found both the genetic correlation of our sex-stratified meta-analysis and the correlation of the male and female effect sizes of SNPs known to be associated with MDD to be significantly below one. However, we benchmarked all our male-female comparisons with male-male and female-female comparisons which demonstrated these results could be due to the inherent heterogeneity of MDD rather than sex differences. A genetic correlation not significantly different to one was found by Blokland *et al.* [16] but not by Silveira *et al.* [17]. The discrepancy between our genetic correlation results (not indicating sex differences) and polygenic overlap results (providing evidence for a set of female-specific SNPs) can be explained by the differing methods. Genetic correlation captures the average effect and direction of pleiotropy across the entire genome, but does not capture variation in correlation at specific loci. Thus, the genetic correlation estimate can be unaffected by a set of trait-specific variants if the majority of variants are near identical in effect size and direction (as is the case here where shared male-female variants were correlated at one) [29]. Overall, these results suggest that the greater genetic contribution to MDD risk in females may not be due to effect size differences but rather due to a set of female-specific SNPs, i.e. our results do not support the sex-dependent hypothesis but do support the hypothesis that sex-specific genetic effects contribute towards sex differences in MDD.

Despite these similarities, we found very little overlap in genes associated with MDD across the sexes. Our gene-based tests and annotation of our genome-wide significant SNPs to genes both identified only one gene significantly associated with MDD in females and males; *DCC* and *NEGR1*, respectively. Both of these genes are involved in neuronal connectivity and genetic variants in these and related genes have previously been associated with psychiatric disorders including MDD [39–41]. Our sex-stratified GWAS meta-analysis of MDD has differential power and thus our male GWAS may not have had the power to identify the genes associated with MDD in females, while genes detected in males only could be male-specific.

Females show a higher prevalence of atypical and immuno-metabolic depression which are characterised by symptoms such as weight gain, increased appetite and hypersomnia as well as immune-inflammatory pathophysiology, metabolic dysregulation and an increased risk for cardiometabolic diseases [42–44]. Our results estimated a larger genetic correlation and polygenic overlap between MDD in females and metabolic traits (body mass index and metabolic syndrome) than MDD in males with these same traits. Many of the genes we identified as shared by MDD and metabolic traits in females only are associated with neurological disorders such as epilepsy and Huntington’s disease, as well as autism. Our results suggest that sex-specific pleiotropy, and thus sex-specific shared pathophysiological mechanisms, may contribute to the sex differences in metabolic symptoms and comorbidities of people with MDD.

Comorbid substance use and MDD show a range of sex differences. Across studies, alcohol use is consistently higher in males than females with MDD [45]. However, the comorbidity between smoking and MDD does not show consistent sex differences; some studies find smoking is higher in males with MDD and others demonstrate it is higher in females [46]. Our findings show a significantly higher genetic correlation between being a regular smoker and MDD in females, than with MDD in males. This suggests that shared pathophysiological mechanisms may contribute to the comorbidity of smoking and MDD to a larger extent in females. We found a non-significant genetic correlation between MDD in both female sand males with alcohol use (drinks per week). This suggests that the comorbidity of MDD and alcohol use, and its sex difference, may largely be driven by environmental factors rather than shared pathophysiological mechanisms.

The heterogeneity of MDD across sexes may also be influenced by the environment and gene-by-environment interactions. Genetic predisposition, trauma, and their interaction have been shown to influence MDD risk [47–50]. Furthermore, genome-by-trauma interaction effects on MDD were larger in males than females [51]. Interestingly, one of the nominally significant SNPs identified in our genotype-by-sex interaction meta-analysis has previously been associated with MDD with trauma exposure (rs11671136 in LD with rs28573687) [47]. Future work examining whether sex-specific gene-by-environment interactions also contribute to sex differences in MDD will likely be informative. Our sex-stratified summary statistics published here will provide a valuable resource for future analyses.

Our study should be interpreted considering some limitations. Firstly, the genome-wide meta-analysis of MDD has a 1.65-fold larger effective sample size in females compared to males (n_eff (Females)_ = 287,082; n_eff (Males)_ = 173,943) and this power difference could exaggerate sex differences identified. Secondly, our analyses are restricted to Europeans only limiting the applicability of our findings to other populations. Thirdly, as the software used to conduct our genotype-by-sex interaction analysis only includes the genotype-by-sex interaction term, and cannot include all genotype-by-covariate and sex-by-covariate interaction terms, confounders may not have been properly controlled for in this analysis [52].

Here, we conducted the largest sex-stratified genome wide association and genotype-by-sex interaction meta-analyses for MDD to date. Our findings reveal a novel genetic variant associated with MDD in males, and provide new evidence for a higher burden of genetic risk in females. We highlight the potential role of the X chromosome in modulating MDD risk differently between sexes. These insights enhance our understanding of the genetic basis for sex differences in MDD and underscore the importance of sex-stratified approaches in genetic research. Moving forward, integrating sex-specific genetic findings into clinical practice could pave the way for more personalized diagnostic and therapeutic strategies for MDD.

## Methods

### Participants

Data from five international cohorts were analysed: The Australian Genetics of Depression Study (AGDS) in Australia [53] (Female: cases=10,406, controls=7,147; Male: cases=3,174, controls=6,601), The BIObanks Netherlands Internet Collaboration (BIONIC) in the Netherlands [54] (Female: cases=10,664, controls=26,878; Male: cases=4,432, controls=20,013), the Genetic Links to Anxiety and Depression (GLAD+) study in the United Kingdom (UK) [55] (Female: cases=16,708, controls=3,393; Male: cases=4,656, controls=3,025), the UK Biobank from the UK [56] (Female: cases=46,194, controls=53,211; Male: cases=22,608, controls=56,516) and All Of Us from the United States of America [57] (Female: cases=26,776, controls=46,678; Male: cases=11,136, controls=35,836). Across all cohorts, MDD cases and controls were defined primarily based on the DSM (Diagnostic and Statistical Manual of Mental Disorders) where available, supplemented by electronic health records and/or self-report of diagnosis (Supplementary Table 1). See Supplementary Methods A for more details on each cohort.

### Genotyping, Quality Control and Imputation

Details about genotyping platform used as well as quality control and imputation methods can be found in Supplementary Methods A. In all cohorts, individual quality control included exclusion of participants with non-European ancestry.

### Association Analyses

All association analyses in all cohorts were conducted using fastGWA in GCTA v1.94.1 [58]. Sex-stratified genome-wide association studies (GWAS) were carried out in each cohort on the autosomes and X chromosome using a mixed linear model for a binary outcome (--fastGWA-mlm-binary) with a sparse genetic relationship matrix and the first 10 ancestry principal components, as well as any cohort-specific covariates. Genome-wide genotype-by-sex interaction (GxS) analyses were also carried out in each cohort using a mixed linear model (--fastGWA-mlm) with a sparse genetic relationship matrix, sex as the environment variable and the first 10 ancestry principal components and any cohort-specific covariates. GxS analyses were conducted for the autosomes, as well as for the X chromosome with the non-pseudoautosomal region analysed both with no dosage compensation (--dc 0) and full dosage compensation (--dc 1). The sex-stratified GWAS and GxS results were filtered to retain SNPs with a minor allele frequency larger than 0.01 and a R^2^ imputation score larger than 0.6. See Supplementary Methods A for cohort specific details.

### Meta-analysis

The sex-stratified GWAS and GxS results from the five cohorts were meta-analysed with summary statistics from Blokland *et al.* [16] which resulted in a total sample size of 130,471 cases and 159,521 controls in females, and 64,805 cases and 132,185 controls in males. As the GxS association analyses in all studies used linear mixed models the beta values were converted to the logistic scale by firstly converting the beta values to odds ratios (OR) using the R function from Lloyd-Jones *et al.* [59] and then using the log of the OR. The standard error of the log(OR) was calculated as the log(OR) divided by the Z-score. Standard error weighted meta-analyses without genomic control were run in METAL [60]. The meta-analysis results were filtered to only retain SNPs present in three or more cohorts (at least 50% of the cohorts). In order to identify lead, independent SNPs, clumping was carried out in PLINK v1.90b6.8 [61, 62] with a linkage disequilibrium threshold (--clump-r2) of 0.1 and a physical distance threshold (--clump-kb) of 1000.

### Replication Cohort

Generation Scotland served as a replication cohort [63]. Case status was determined by case classification in the Structured Clinical Interview for DSM-IV disorders (SCID) [64, 65] or the Composite International Diagnostic Interview (CIDI) [66]. Individuals also present within the UK Biobank and of non-European ancestry were removed, resulting in a sample size of 2,441 cases and 3,321 controls in females, and 938 cases and 2,491 controls in males. Sex-stratified association analyses were run using fastGWA in GCTA v1.94.1 [58] (--fastGWA-mlm-binary) with a sparse genetic relationship matrix and the first four ancestry principal components as covariates. We compared the beta-values from the lead independent genome-wide significant SNPs from our meta-analysis to the replication analysis. For females all 16 lead SNPs were tested and seven lead SNPs were tested in males (one SNP was unavailable in the Generation Scotland cohort). A binomial test was used to determine whether significantly more than 50% of the SNPs tested in the replication analysis (expected by chance) had a concordant effect size direction. See Supplementary Methods A for more details on the Generation Scotland cohort.

### GxS Sensitivity Analyses

As interaction analyses require very high power in order to detect significant interactions, we also carried out a sensitivity analysis to determine whether any SNPs known to be associated with sex-combined MDD had a significant GxS interaction. We used the genome-wide significant SNPs from the largest MDD GWAS meta-analysis by Adams *et al.* [27] and a P-value threshold of 7.17 x 10^-5^ (0.05/697 independent SNPs).

### Genetic Architecture of MDD in Females and Males

Autosomal SNP-based heritability (*h^2^*_SNP_), polygenicity (π) and the selection parameter (*S*), a measure of the relationship between effect size and minor allele frequency which indicates the direction and strength of natural selection, were estimated using our sex-stratified meta-analysis summary statistics in SBayesS using GCTB v2.5.2 [67]. The linkage disequilibrium shrunk sparse matrix of 2.8 million variants [68] and four chains of length 25,000 with a burn-in of 5,000 and thinning of 10 were used. *H^2^*_SNP_ was converted to the liability scale using the method from Lee *et al.* [69] with a population prevalence of 0.2 and 0.1 in females and males, respectively. To determine whether *h^2^*_SNP_, π and *S* are different across sexes, we calculated the posterior probability that female value > male value by counting the frequency of Markov chain Monte Carlo (MCMC) samples in which female value > male value. We conducted a range of sensitivity analyses comparing the genetic architecture of MDD in females and males (Supplementary Methods B). This included calculating *h^2^*_SNP_ estimates in LDSC, accounting for the differential power of our sex-stratified GWAS meta-analysis, as well as examining the role of male under-diagnosis and across-cohort heterogeneity.

### Genetic Correlation

Linkage Disequilibrium Score Regression (LDSC) [70, 71] with European LD scores computed from 1000 Genomes was used to estimate bivariate autosomal SNP-based genetic correlations (*r_g_*). We estimated *r_g_* between our sex-stratified summary statistics with the largest GWAS meta-analysis of sex-combined MDD (including 23andMe, Inc and Europeans only) [27] and previous sex-stratified GWAS of MDD [16, 17]. We also estimated *r_g_* between our male and female meta-analysis summary statistics. To account for across-cohort heterogeneity we also estimated *r_g_* between each pairwise combination of the two sexes by six cohorts (the five new cohorts and the existing GWAS meta-analysis from Blokland *et al.* [16] used in our meta-analyses here).

We also investigated whether the MDD effect sizes (betas) for SNPs known to be associated with sex-combined MDD are different across the sexes. We entered all of the genome-wide significant hits associated with MDD from Adams *et al.* [27] (Europeans only) into PLINK v1.90b6.8 [61, 62] clumping with a linkage disequilibrium threshold (--clump-r2) of 0.1 and a physical distance threshold (--clump-kb) of 1000 to identify lead, independent SNPs. For these SNPs, ensuring the minor allele was the tested allele and consistent across summary statistics, the Pearson correlation was calculated between the standardised effect size estimates (beta values) of our male and female meta-analysis summary statistics, and between each pairwise combination of the two sexes by six studies. Beta values and their standard errors were standardised accounting for the influence of allele frequency and sample size (Formula 1 and Formula 2), where Z represents the Z statistic, p the allele frequency and n the sample size.

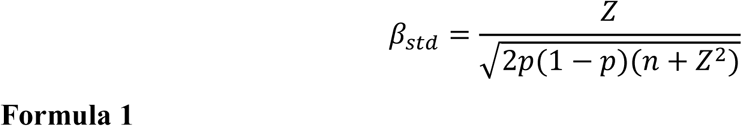

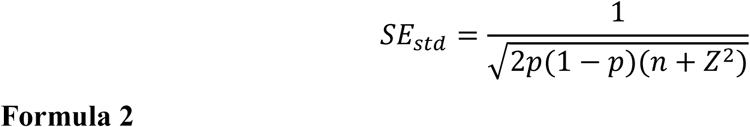

For both the *r_g_* estimates and Pearson’s correlations, an inverse-variance weighted meta-analysis using a random-effects model (restricted maximum likelihood estimator) in R v4.3.1 [72] and the ‘metafor’ package [73] was completed for the six male-female comparisons within studies, the 30 male-female comparisons across studies, the 15 male-male comparisons across studies and the 15 female-female comparisons across studies (same sex within study comparisons were not included as they would yield a correlation of 1) (Supplementary Figures 4 – 6). Within-sex comparisons were included as a baseline check to determine whether male-female differences were due to across-sex or across-study heterogeneity. To determine whether *r_g_* estimates / Pearson correlations were significantly different from one we calculated the Z-score, where *H_o_* = the null hypothesis (1 in this case) (Formula 3). The P-value was calculated from a standard normal distribution using a two-tailed test and adjusted for five comparisons using the Benjamini-Hochberg method.

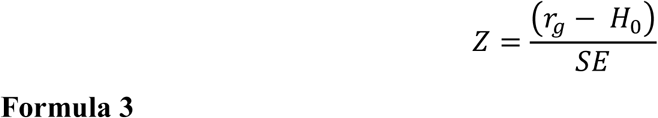

A similar approach was repeated using the slope and intercept from linear regressions of male versus female effect size estimates (see Supplementary Methods C).

### Polygenic Overlap of MDD Across Sexes

We used univariate and bivariate MiXeR v1.3 [28, 29] to quantify polygenicity of male and female MDD and estimate polygenic overlap between sexes. MiXeR fits a Gaussian mixture model assuming that common genetic effects on a trait are a mixture of causal variants and non-causal variants. Polygenicity is reported as the number of causal variants that explain 90% of *h^2^_SNP_* (to avoid extrapolating model parameters into the area of infinitesimally small effects). As a sensitivity analysis to account for the differential power of our sex-stratified GWAS, we also ran univariate and bivariate MiXeR on the full UK Biobank sample (n_Females_ = 46,194 cases and 53,211 controls, n_Males_ = 22,608 cases and 56,516 controls) and after down-sampling (n = 22,608 cases and 53,211 in both females and males).

We used gwas-pw [30] to identify causal risk loci for MDD that are sex-specific or shared across the sexes. The recommended European bed file that splits the genome (autosomes only) into approximately independent LD blocks was used within gwas-pw and correlation was set as zero because our female and male MDD GWAS do not have any overlapping individuals. Genomic regions with a posterior probability of association (PPA) larger than 0.5 for model one, two and three were identified as regions containing a causal variant for MDD in females only, males only or shared by both sexes, respectively. Subsequently, for each of these identified regions the SNP with the largest PPA that was above 0.5 was selected as a possible causal MDD variant. These possible causal variants were then annotated using SNP2GENE from FUMA v1.5.2 [32] with ANNOVAR (2017-07-17) and ensemble v110.

Positional mapping was carried out with a window size of 10kb, eQTL mapping using all the tissue types from TIGER [74], InsPIRE [75], EyeGEx [76], eQTL catalogue, PsychENCODE [77], van der Wijst *et al.* [78] scRNA eQTLs, DICE [79], eQTLGen, Blood eQTLs [80], MuTHER [81], xQTLServer [82], CommonMind Consortium [83], BRAINEAC [84] and GTEx v8 [85], and 3D Chromatin Interaction Mapping using all Buildin chromatin interaction data (Hi-C of 21 tissue/cell types from GSE87112 [86], Hi-C loops from Giusti-Rodríguez *et al.* [87], Hi-C based data from PsychENCODE [77] and Enhancer-Promoter correlations from FANTOM5 [88–90]). All of the annotation datasets from PsychENCODE, FANTOM5 and Brain Open Chromatin Atlas [91] were used for all three mapping types. Genes annotated to a SNP with more than one method were considered.

### Functional annotation and analyses

Gene-based, gene-set and gene-property tests were carried out for the sex-stratified and GxS results using MAGMA v1.08 [31] within the SNP2GENE function in FUMA v1.5.2 [32]. Our summary statistics as well as the lead, independent SNPs as determined by PLINK clumping were used as inputs. Settings used were ensembl v110, ANNOVAR (2017-07-17) [92], maximum P-value of lead SNPs and P-value cut-off of 5 x 10^-8^ (sex-stratified summary statistics) or 1 x 10^-6^ (GxS summary statistics), a first and second R^2^ threshold of 0.1, the European 1000Genomes Phase 3 reference panel [93], and a gene window size of 10 kb. The gene-based test used a genome-wide significance P-value of 2.53 x 10^-6^ as the input SNPs mapped to 19,759 protein coding genes. The gene-set analysis used 10,678 gene sets (4,761 curated gene sets and 5,917 Gene Ontology terms) from MsigDB v6.2 [94] and significance was determined using Bonferroni corrected P-values. Gene property analysis for tissue specificity was conducted using 30 general tissue types and 53 tissue types from GTEx v8 [85]. Genome-wide significant SNPs (sex-stratified analysis) and nominally significant SNPs (GxS analysis) underwent gene annotation using positional, eQTL and 3D Chromatin Interaction mapping using the same settings as specified above (***‘Polygenic Overlap of MDD Across Sexes’***).

To determine whether any of our genome-wide significant SNPs (sex-stratified analysis) and nominally significant SNPs (GxS analysis) are novel and to search for previous SNP-phenotype associations, a second analysis using the SNP2GENE function in FUMA v1.5.2 with GWASCatalog (e0_r2022-11-29) was run. The settings as described above were used, however the maximum P-value cut-off was changed to one, and SNPs from the reference panel were included. A lead independent significant SNP was considered novel if the significant SNP, or any SNPs in linkage disequilibrium with it, had not previously been associated with any depression phenotypes according to the list of SNPs in GWASCatalog. As the results from the largest GWAS meta-analysis of MDD [27] had not yet been added to GWASCatalog at the time of our analyses, all significant SNPs, and those in LD with them, were also checked against all of the genome-wide significant SNPs, and any SNPs in LD, in this publication. SNPs in LD were determined using PLINK v1.90b6.8 [61, 62] with a linkage disequilibrium threshold (--clump-r2) of 0.1 and a physical distance threshold (--clump-kb) of 1000.

### Sex-specific pleiotropic effects

We used LDSC to estimate genome-wide autosomal SNP-based *r_g_* between our sex-stratified MDD GWAS meta-analysis results and a range of other psychiatric disorders, metabolic traits and substance use traits (Supplementary Table 21). Metabolic and substance use traits were included to determine whether sex-specific pleiotropic effects may contribute to sex differences in metabolic symptoms and substance use in people with MDD. As body mass index (BMI) has a well-powered sex-stratified GWAS [95], we also estimated *r_g_* for all bivariate combinations of our sex-stratified MDD GWAS and the sex-stratified BMI summary statistics.

To determine whether *r_g_* between MDD and each trait is significantly different across sexes we used the jack-knife method, a resampling technique which systematically leaves out one block of data at a time. Unlike the Z-score method, which is commonly used, the jack-knife method accounts for LD structure and does not assume independence of the two genetic correlation values, which is important as the same second trait is used in both the genetic correlations being compared (e.g. MDD in females versus ADHD compared to MDD in males versus ADHD). In LDSC, the –-print-delete-vals flag was used to obtain delete values of genetic covariance and heritabilities for 200 blocks. For each bivariate genetic correlation, delete values of genetic correlation were calculated for each of the 200 blocks (**Error! Reference source not found.**), where G = genetic covariance and *h^2^* = SNP-based heritability.

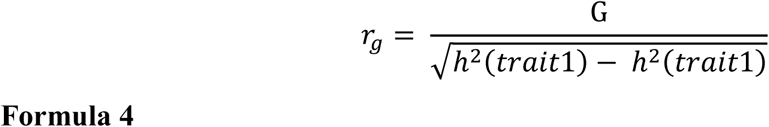

Jack-knife pseudo-values were then calculated for each of the 200 blocks (Formula 5), where P = jack-knife pseudo-value, n = total number of jack-knife blocks (200 in this case), 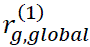 = the global genetic correlation obtained from LDSC for comparison one, 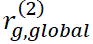 = the global genetic correlation obtained from LDSC for comparison two, 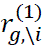 = the delete genetic correlation estimated with block *i* removed from comparison one, and 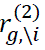 = the delete genetic correlation estimated with block *i* removed from comparison two.

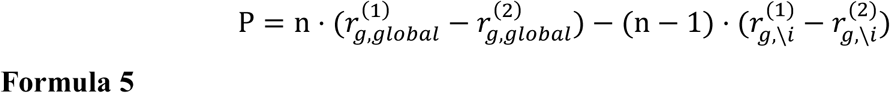

The mean and standard error of the 200 jack-knife pseudo-values estimates the difference between the two genetic correlations and was compared to the null hypothesis that there is no difference between the genetic correlations by calculating the Z-score, where *H_o_* = the null hypothesis (0 in this case) (Formula 3). The P-value was calculated from a standard normal distribution using a two-tailed test followed by the Benjamini Hochberg correction for 11 tests (all traits) or four tests (sex-specific BMI). As a sensitivity analysis, the Z-score method was also used to determine whether *r_g_* between MDD and each trait is significantly different across sexes because although it is not theoretically appropriate it is commonly used (Supplementary Methods D).

We further explored the metabolic traits BMI and metabolic syndrome that showed significant sex-specific *r_g_*. To quantify polygenic overlap between MDD in females/males and each metabolic trait, we used MiXeR v1.3 [28, 29]. Gwas-pw [30] was used to identify causal risk loci shared by MDD and each metabolic trait that are sex-specific or shared across the sexes. Inputs to gwas-pw were our female MDD summary statistics and female BMI summary statistics [95], or our male MDD summary statistics and male BMI summary statistics [95], and the recommended European PLINK bed file that splits the genome (autosomes only) into approximately independent LD blocks. The correlation between beta values of SNPs within genomic regions with a PPA < 0.2 as calculated in fgwas [96] was used to account for potential overlapping of cohorts. Genomic regions with a PPA > 0.5 for model 3 were compared between the female MDD – female BMI and male MDD – male BMI analyses to identify genomic regions with a causal variant shared by MDD and BMI that are female-specific, male-specific or shared by both sexes. Subsequently, for each of these identified regions the SNP with the largest PPA that was above 0.5 was selected as a causal MDD/BMI variant. These causal variants were then annotated using SNP2GENE from FUMA v1.5.2 [32] with the same settings as described above (***‘Polygenic Overlap of MDD Across Sexes’***). The above was repeated for the comparison of the sex-combined metabolic syndrome summary statistics [97] with our female and male MDD summary statistics.

## Acknowledgements

We would like to thank the research participants from each of the cohorts for giving their time to make this work possible. We also thank the employees from All Of Us and UK Biobank.

## The Australian Genetics of Depression Study (AGDS)

AGDS was primarily funded by the National Health and Medical Research Council (NHMRC) of Australia Grant No. 1086683, and the QSkin study was funded by the NHMRC (Grant Numbers 1185416, 1063061 and 1073898). We thank everyone who contributed to the conception, implementation, media campaign and data cleaning of the AGDS project, including Richard Parker, Simone Cross, and Lenore Sullivan. Thank you to Scott Gordon for carrying out the imputation and quality control of the AGDS and QSkin data and Penelope Lind for contributing to the management of the AGDS database. This work was supported by NHMRC Investigator Grants to Jackson G Thorp (2027002); Brittany L Mitchel (2017176); Sarah E Medland (1172917); Nicholas G Martin (1172990); NRW (1173790); David C. Whiteman (2026567) and Ian B Hickie (2016346). Enda M Byrne received funding from the NHMRC-funded PRE-EMPT Centre of Research Excellence.

## UK Biobank

The UK Biobank Resource (application number 25331) was used for this work.

## BIONIC

To create the BIONIC consortium we are grateful to for funding from the Biobanking and Biomolecular Resources Research Infrastructure (BBMRI-NL: 184.021.007; 184.033.111) to Dorret I Boomsma and Brenda WJH Penninx. Floris Huider was funded by the KNAW Academy Professor Award (PAH/6635) to Dorret I Boomsma. Hanna M van Loo was supported by a NARSAD Young Investigator grant and by National Institute of Mental Health (NIMH) grant R01MH125902.

## BIONIC consortium authors

Floris Huider^1,2,3^, Yuri Milaneschi^2,4,5^, René Pool^1,2^, M. Liset Rietman^6^, Almar A.L. Kok^2,4,7^, Tessel E. Galesloot^8^, Leen M. ‘t Hart^2,7,9,10^, Femke Rutters^2,7^, Marieke T. Blom^2,11^, Didi Rhebergen^2,4,5^, Marjolein Visser^2,12^, Ingeborg A. Brouwer^2,12^, Edith Feskens^13^, Catharina A. Hartman^14^, Albertine J. Oldehinkel^14^, Mariska Bot^2,4,5^, Eco J.C. de Geus^1,2^, Lambertus A. Kiemeney^8,15^, Martijn Huisman^2,7,16^, H. Susan J. Picavet^6^, W.M. Monique Verschuren^6,17^, Hanna M. van Loo^14^, Brenda W.J.H. Penninx^2,4,5^, Jouke-Jan Hottenga^1,2,18^, Dorret I. Boomsma^2,3^

1. Department of Biological Psychology, Faculty of Behavioral and Movement Sciences, Vrije Universiteit Amsterdam, 1081 Amsterdam, The Netherlands.
2. Amsterdam Public Health Research Institute, Amsterdam, The Netherlands.
3. Department of Complex Trait Genetics, Vrije Universiteit Amsterdam, 1081 Amsterdam, The Netherlands.
4. Department of Psychiatry, Amsterdam UMC location Vrije Universiteit Amsterdam, 1081 Amsterdam, The Netherlands.
5. Amsterdam Neuroscience, Complex Trait Genetics, 1081 Amsterdam, The Netherlands.
6. Center for Prevention, Lifestyle and Health, Dutch National Institute for Public Health and the Environment, 3721 Bilthoven, The Netherlands.
7. Department of Epidemiology and Data Science, Amsterdam UMC, 1081 Amsterdam, The Netherlands.
8. IQ Health Science Department, Radboud University Medical Center, 6525 Nijmegen, The Netherlands.
9. Department of Cell and Chemical Biology, Leiden University Medical Center, 2333 Leiden, The Netherlands.
10. Department of Biomedical Data Sciences, Section Molecular Epidemiology, Leiden University Medical Center, 2333 ZA Leiden, The Netherlands.
11. Department of General Practice, Amsterdam UMC, 1081 Amsterdam, The Netherlands.
12. Department of Health Sciences, Faculty of Science, Vrije Universiteit Amsterdam, 1081 Amsterdam, The Netherlands.
13. Division of Human Nutrition and Health, Wageningen University & Research, 6700 Wageningen, The Netherlands.
14. Department of Psychiatry, University of Groningen, University Medical Center Groningen, 9713 Groningen, The Netherlands.
15. Department of Urology and Department for Health Evidence, Radboud University Medical Center, 6525 Nijmegen, The Netherlands
16. Department of Sociology, Vrije Universiteit Amsterdam, 1081 Amsterdam, The Netherlands.
17. Julius Center for Health Sciences and Primary Care, University Medical Center Utrecht, 3584 Utrecht, The Netherlands.
18. Neurological Disorder Research Center, Qatar Biomedical Research Institute (QBRI), Hamad Bin Khalifa University (HBKU), Qatar Foundation, Doha P.O. Box 5825, Qatar.

## The GLAD Study Group authors

Gursharan Kalsi^1,2^, Saakshi Kakar^1,2^, Christopher Hübel^1,2,3,4^, Ian Marsh^1,2^, Laura H Meldrum^1,2^, Iona Smith^1,2^, Jahnavi Arora^1,2^, Henry C. Rogers^1,2,5^, Brett N. Adey^1,2^, Zain Ahmad^1^, Shannon Bristow^1,2^, Charles J. Curtis^1,2^, Susannah C. B. Curzons^1,2^, Helena L. Davies^1,6,7^, Molly R. Davies^8^, Abigail R. ter Kuile^1,2^, Sang Hyuck Lee^1,2^, Yuhao Lin^1,2^, Jared G. Maina^1,2^, Monika McAtarsney-Kovacs^1,2^, Dina Monssen^1,2^, Jessica Mundy^1,2^, Alish B. Palmos^1,2^, Alicia J. Peel^1,2^, Kirstin Purves^1,2^, Christopher Rayner^1,2^, Megan Skelton^1,2^, Katherine N. Thompson^1,9^, Rujia Wang^1,2^, Johan Zvrskovec^1,2^, Joshua E. J. Buckman^10^, Ewan Carr^11^, Antony J. Cleare^12,13^, Katrina A. S. Davis^8^, Kimberly A. Goldsmith^11^, Colette R. Hirsch^14,15^, Georgina Krebs^1,16^, Donald M. Lyall^17^, Katharine A. Rimes^12^, Evangelos Vassos^1,2^, David Veale^12,13^, Janet Wingrove^18^, Allan H. Young^19^, Roland Zahn^19^, Le Roy Dowey^20^, Victor Gault^20^, Chérie Armour^21^, John R. Bradley^22^, Ian R. Jones^23^, Nathalie Kingston^22^, Andrew M. McIntosh^24^, Daniel J. Smith^25^, James T. R. Walters^26^, NIHR BioResource consortium, Jonathan R. I. Coleman^1,2^, Matthew Hotopf^2,8^, Thalia C. Eley^1,2^, Gerome Breen^1,2^

1. Social, Genetic, and Developmental Psychiatry Centre; Institute of Psychiatry, Psychology and Neuroscience; King’s College London, London, UK
2. UK National Institute for Health Research (NIHR) Biomedical Research Centre, South London and Maudsley Hospital and King’s College London, London, UK
3. National Centre for Register-based Research, Aarhus University, Aarhus, Denmark
4. Department of Pediatric Neurology, Charité – Universitätsmedizin Berlin, Berlin, Germany
5. Department of Psychiatry, Mount Sinai Health System, New York, USA
6. Mental Health Center Ballerup, Copenhagen University Hospital – Mental Health Services CPH, Center for Eating and feeding Disorders Research, Copenhagen, Denmark
7. Institute of Biological Psychiatry, Mental Health Center Sct. Hans, Mental Health Services Copenhagen, Roskilde, Denmark
8. Department of Psychological Medicine, Institute of Psychiatry, Psychology & Neuroscience, King’s College London, London, UK
9. Department of Sociology, College of Liberal Arts, Purdue University, West Lafayette, IN, USA
10. CORE Data Lab, Centre for Outcomes Research and Effectiveness (CORE), Research Department of Clinical, Educational, and Health Psychology, UCL, London, UK
11. Department of Biostatistics and Health Informatics, Institute of Psychiatry, Psychology and Neuroscience, King’s College London, London, UK
12. The Institute of Psychiatry, Psychology and Neuroscience, King’s College London, London, UK
13. South London and Maudsley NHS Foundation Trust, Maudsley Hospital, London, UK
14. Department of Psychology, Institute of Psychiatry, Psychology and Neuroscience, King’s College London, Denmark Hill, Camberwell, London, UK
15. Centre for Anxiety Disorders and Trauma, South London and Maudsley Hospital, London, UK
16. Research Department of Clinical, Educational and Health Psychology, University College London, London, UK
17. Institute of Health and Wellbeing, University of Glasgow, Glasgow, UK
18. Talking Therapies Southwark, South London and Maudsley NHS Foundation Trust, London, UK
19. South London and Maudsley NHS Foundation Trust and Centre for Affective Disorders, Department of Psychological Medicine, Institute of Psychiatry, Psychology, and Neuroscience, King’s College London, London, UK
20. School of Biomedical Sciences, Ulster University, Coleraine, Northern Ireland, UK
21. Research Centre for Stress Trauma and Related Conditions (STARC), School of Psychology, Queen’s University Belfast, Belfast, UK
22. University of Cambridge, Cambridge, UK
23. National Centre for Mental Health, Cardiff University, Cardiff, UK
24. Division of Psychiatry, Centre for Clinical Brain Sciences, University of Edinburgh, Edinburgh, UK
25. Division of Psychiatry, Centre for Clinical Brain Sciences, University of Edinburgh, Royal Edinburgh Hospital, Edinburgh, UK
26. Centre for Neuropsychiatric Genetics and Genomics, Division of Psychological Medicine and Clinical Neurosciences, School of Medicine, Cardiff University, Cardiff, UK

## Generation Scotland

Generation Scotland received core support from the Chief Scientist Office of the Scottish Government Health Directorates (CZD/16/6) and the Scottish Funding Council (HR03006) and is currently supported by the Wellcome Trust (216767/Z/19/Z). Genotyping of the Generation Scotland samples was carried out by the Genetics Core Laboratory at the Edinburgh Clinical Research Facility, University of Edinburgh, Scotland and was funded by the Medical Research Council UK and the Wellcome Trust (Wellcome Trust Strategic Award “STratifying Resilience and Depression Longitudinally” (STRADL) Reference 104036/Z/14/Z).

The genome-wide summary statistics for the Adams *et al.* [27] analysis of 23 andMe, Inc., data were obtained under a data transfer agreement with QIMR Berghofer. We would like to thank the research participants and employees of 23andMe, Inc. for making this work possible.

## Data Availability

All GWAS summary statistics are available from GWAS Catalog (accession numbers GCST90565869, GCST90565870, GCST9056587, and GCST90565872) (embargoed until publication) and from the authors upon request.

## Code Availability

Code used to conduct analyses presented in this manuscript is available from Github (https://github.com/jodithea/Sex_differences_genetics_depression) and has been archived on Zenodo and assigned a DOI (https://doi.org/10.5281/zenodo.15233099) (embargoed until publication).

## Author contributions

J.T. Thomas, N.G. Martin and B.L. Mitchell conceived the study. Collection, contribution of data and/or supervision of analysts was provided by the following people for each cohort: AGDS = N.G. Martin, S.E. Medland, I.B. Hickie, E.M. Byrne, N.R. Wray, B.L. Mitchell; QSkin = D.C. Whiteman, C.M. Olsen; All Of Us = E.M. Derks, Bionic = D.I. Boomsma, B.W.J.H. Penninx, H.M. van Loo; GLAD+ = J.R.I. Coleman, T.C. Eley, G. Breen, Generation Scotland = M. Adams, H.C. Whalley. Analyses in each cohort were carried out as follows: AGDS = J.T. Thomas, All Of Us = J.G. Thorp and P. Youssef, Bionic = F. Huider, GLAD+ = R. Wang, Generation Scotland = P.Z. Grimes, UK Biobank = J.T. Thomas. J.T. Thomas performed the meta-analyses and most of the downstream statistical and bioinformatics analyses, with MiXeR analyses being carried out by J.G. Thorp. Supervision throughout the project was provided by B.L. Mitchell. Support and advice with statistical analyses and interpretation was provided by N.R. Wray and J.R.I. Coleman. J.T. Thomas, B.L. Mitchell and J.G. Thorp wrote the manuscript, with all authors providing comments and suggestions.

## Competing Interests

The authors declare no competing interests.

## Notes

### Competing Interest Statement

The authors have declared no competing interest.

### Funding Statement

The Australian Genetics of Depression Study (AGDS) was primarily funded by the National Health and Medical Research Council (NHMRC) of Australia Grant No. 1086683, and the QSkin study was funded by the NHMRC (Grant Numbers 1185416, 1063061 and 1073898). This work was supported by NHMRC Investigator Grants to Jackson G Thorp (2027002); Brittany L Mitchel (2017176); Sarah E Medland (1172917); Nicholas G Martin (1172990); NRW (1173790); David C. Whiteman (2026567) and Ian B Hickie (2016346). Enda M Byrne received funding from the NHMRC-funded PRE-EMPT Centre of Research Excellence.
To create the BIONIC consortium we are grateful to for funding from the Biobanking and Biomolecular Resources Research Infrastructure (BBMRI-NL: 184.021.007; 184.033.111) to Dorret I Boomsma and Brenda WJH Penninx. Floris Huider was funded by the KNAW Academy Professor Award (PAH/6635) to Dorret I Boomsma. Hanna M van Loo was supported by a NARSAD Young Investigator grant and by National Institute of Mental Health (NIMH) grant R01MH125902.
Generation Scotland received core support from the Chief Scientist Office of the Scottish Government Health Directorates (CZD/16/6) and the Scottish Funding Council (HR03006) and is currently supported by the Wellcome Trust (216767/Z/19/Z). Genotyping of the Generation Scotland samples was funded by the Medical Research Council UK and the Wellcome Trust (Wellcome Trust Strategic Award 'STratifying Resilience and Depression Longitudinally' (STRADL) Reference 104036/Z/14/Z).

### Author Declarations

Ethics committee of QIMR Berghofer Medical Research Institute gave ethical approval for this work(P2118, P1309 and P2034).

