## Supplementary File for "Sex-stratified genome-wide association meta-analysis of Major Depressive Disorder"

**Supplementary Results A:**

**Sensitivity Analyses for the Genetic Architecture of Depression in Females and Males**

***LDSC***

When using LDSC with our sex-stratified meta-analysis results, $h^2_{SNP}$ estimates on the liability scale were significantly higher in females (9.7 ± 0.4%, $h^2$ ± SE) compared to males (7.8 ± 0.5%) (H0: $h^2_F - h^2_M = 1$ [Z = 2.97, p = 0.003]), when using a population prevalence of 0.2 in females and 0.1 in males.

***Differential Power***

Our female GWAS has a 1.65-fold larger effective sample size compared to that in males ($n_{eff\ (Females)}$ = 287,082; $n_{eff\ (Males)}$ = 173,943). To test whether our results were influenced by this power difference we ran SBayesS on the full UK Biobank sample ($n_{Females}$ = 46,194 cases and 53,211 controls, $n_{Males}$ = 22,608 cases and 56,516 controls) and after down-sampling (n = 22,608 cases and 53,211 in both females and males). In both the full and down-sampled UK Biobank we found very strong evidence (full = 99% posterior probability (PP), down-sampled = 100% PP) that $h^2_{SNP}$ estimates on the liability scale are higher in females than males (Full: $h^2_{SNP}$ female = 14.9% [95% highest posterior density interval (HPDI): 13.5 – 16.2%]; $h^2_{SNP}$ male = 12.7% [11.1 – 14.4%]. Down-sampled: $h^2_{SNP}$ female = 17.1% [14.8 – 19.2%]; $h^2_{SNP}$ male = 12.3% [10.4 – 13.9%]) (Supplementary Figure 3A). In the full UK Biobank there was moderate evidence (87% PP) that polygenicity is higher in females ($\pi$ = 0.013 [0.009 – 0.017]) than males ($\pi$ = 0.011 [0.007 – 0.015]). In the down-sampled UK Biobank there was evidence against (29% PP) the polygenicity being higher in females ($\pi$ = 0.010 [0.007 – 0.013]) than males ($\pi$ = 0.012 [0.006 – 0.018]) (Supplementary Figure 3B). These sensitivity analyses suggest $h^2_{SNP}$ is higher in females than males even when accounting for the power difference across sexes. The result of MDD in females being more polygenic than MDD in males could be driven by the power imbalance of our sex-stratified GWAS. However, estimation of polygenicity generally requires more GWAS power than that of heritability and the female and male polygenicity estimates in both the full and down-sampled UK Biobank samples show largely overlapping 95% highest posterior density intervals. This suggests that we may not have enough power in our UK Biobank sample to test whether the difference in polygenicity estimates across sexes is due to a power imbalance. As the effect of differential power on the SBayesS polygenicity estimate was inconclusive we also applied univariate MiXeR to the full and down-sampled UK Biobank cohort. The number of causal variants explaining 90% of MDD $h^2_{SNP}$ was estimated to be 11,242 (SE = 1,366) for females and 7,620 (SE = 2,091) for males in the full UK Biobank, and 12,414 (SE = 4,077) for females and 6,777 (SE = 1,514) for males in the down-sampled UK Biobank (Supplementary Table 9). These results suggest MDD is more polygenic in females than males even when accounting for the power difference across sexes.

#### *Male Under-diagnosis*

When the same population prevalence is used for males and females, we found that $h^2_{\mathrm{SNP}}$ on the liability scale is similar across sexes, suggesting the sex difference in $h^2_{\mathrm{SNP}}$ is driven by the prevalence difference (Supplementary Figure 3G). For common disorders, such as MDD, $h^2_{\mathrm{SNP}}$ on the liability scale can be underestimated when the controls are not screened, i.e. the controls are contaminated with cases [1]. As some studies report that MDD in males is under-diagnosed [2] we estimated $h^2_{\mathrm{SNP}}$ on the liability scale for males accounting for unscreened controls and a corresponding increase in population prevalence. Assuming MDD in females has no unscreened controls and a population prevalence of 0.2, we found at least moderate evidence (> 80% PP) that the estimated $h^2_{\mathrm{SNP}}$ of MDD in females is higher than that in males when males have < 30% unscreened controls and a corresponding population prevalence of < 0.13 (Supplementary Figure 3H) (Supplementary Table 10).

#### *Across-cohort Heterogeneity*

To test whether our results were being driven by across-cohort heterogeneity, we conducted sensitivity analyses in which SBayesS was run in each cohort separately and all Markov chain Monte Carlo (MCMC) samples were combined. When combining the $h^2_{\mathrm{SNp}}$ estimates using SBayesS in each cohort separately, we found strong evidence (93% PP) that $h^2_{\mathrm{SNP}}$ estimates on the liability scale are higher in females ($h^2_{\mathrm{SNP}}$ = 19.4% [4.3 – 47.4%) compared to males ($h^2_{\mathrm{SNP}}$ = 12.5% [0.02 – 35.7%]) using a lifetime population risk of 0.2 and 0.1 in females and males, respectively (Supplementary Figure 3D). When combining the polygenicity ($\pi$) estimates using SBayesS in each study separately, we found moderate evidence (79% PP) that $\pi$ estimates are higher in females ($\pi$ = 0.013 [0.00002 – 0.027]) compared to males ($\pi$ = 0.008 [0.0001 – 0.021]) (Supplementary Figure 3E). When combining the selection parameter ($S$) estimates using SBayesS in each study separately, there was evidence against (24% PP) the selection parameter being higher in females ($S$ = -0.36 [-1.26 – 0.86]) compared to males ($S$ = 0.13 [-1.09 – 2.27]) (Supplementary Figure 3F).

### Supplementary Results B:

#### Male vs Female Linear Regression

Linear regressions were used to investigate whether the MDD effect sizes (beta values) for SNPs known to be associated with sex-combined MDD are different across the sexes (Supplementary Figures 7 - 10). The linear regression slopes of male vs female MDD effect sizes was significantly less than one when using our sex-stratified meta-analysis results (slope = 0.81 ± 0.02, H0: slope = 1, [Z = -9.0, $p_{adj}$(Benjamini-Hochberg) = 2.97 x $10^{-19}$]) and for the meta-analysis of the 30 male-female across cohort comparisons (slope = 0.41 ± 0.04, H0: slope = 1, [Z = -15.19, $p_{adj}$(B-H) = 7.37 x $10^{-52}$]). These estimates are benchmarked against the distribution of male-male (mean slope = 0.31 ± 0.0002 (SE)) and female-female (mean slope = 0.41 ± 0.0002) across cohort comparison meta-analyses. The slope from the meta-analysis of male vs female across cohort comparisons is significantly higher than the distribution of slopes from the meta-analyses of male vs male across cohort comparisons (Z = 2.41, $p_{adj}$(B-H) = 0.03) and not significantly different to the distribution of slopes from the meta-analyses of female vs female across cohort comparisons (Z = -0.18, $p_{adj}$(B-H) = 0.86). Therefore, the male vs female slopes being significantly less than one could be due to the inherent heterogeneity of MDD rather than sex differences in the MDD effect sizes of SNPs associated with sex-combined MDD risk. However, removing cohort variation by conducting a meta-analysis of the slopes from all six female-male within cohort comparisons showed a $r_g$ estimate significantly less that one (slope = 0.44 ± 0.06, H0: slope = 1, [Z = -8.94, $p_{adj}$(B-H) = 3.91 x $10^{-19}$]). This suggests that sex differences in genetic effects that are not fully explained by across cohort heterogeneity may exist within cohorts.

The intercept for male vs female linear regression was not significantly different to 0 when using the sex-stratified meta-analysis results (intercept = -0.00002 ± 0.0002, H0: intercept = 0, [Z = -0.13, $p_{adj}$(B-H) = 0.89]), and the meta-analysis of the male-female within cohort comparisons (intercept = 0.0004 ± 0.0004, H0: intercept = 0, [Z = 1.09, $p_{adj}$(B-H) = 0.34]). The intercept was significantly above 0 for the meta-analysis of the female-male across cohort comparisons (intercept = 0.0005 ± 0.0002, H0: intercept = 0, [Z = 2.68, $p_{adj}$(B-H) = 0.012]). However, the baseline checks show that the intercept from the meta-analysis of male vs female across cohort comparisons is not significantly different to the distribution of intercepts from the meta-analyses of male vs male across cohort comparisons (Z = -0.28, $p_{adj}$(B-H) = 0.78) and to the distribution of intercepts from the meta-analyses of female vs female across cohort comparisons ($Z = 0.71$, $p_{adj}$(B-H) = 0.78). Therefore, there is no evidence that the intercept from the male vs. female linear regression differs from zero, suggesting no systematic shift in the effect sizes of SNPs associated with MDD risk between sexes.

### Supplementary Results C:

#### Genome-wide genotype-by-sex interaction analysis

Two of the four nominally significant independent SNPs identified from the genome-wide genotype-by-sex interaction analyses (GxS) have an effect in opposite directions, with the minor allele increasing MDD risk in females and decreasing risk in males and are located in chromosome 1 (rs12092435, $p = 4.41 \times 10^{-7}$) and chromosome 19 (rs28573687, $p = 6.57 \times 10^{-7}$). The SNP on chromosome 12 (rs12312238, $p = 7.7 \times 10^{-7}$) has no effect in females but decreases MDD risk in males, while the SNP on chromosome 20 (rs6080675, $p = 6.24 \times 10^{-8}$) increases the risk of MDD in both sexes with a larger effect in males (Supplementary Figure 17).

##### *Functional annotation and analyses*

None of the four independent nominally-significant SNPs identified by the GxS analyses, nor any SNPs in linkage disequilibrium with them, have previously been associated with any depression phenotypes according to the list of SNPs in GWASCatalog and the largest MDD GWAS of both sexes combined [3]. Previous associations of these SNPs include metabolic traits such as glucose, lipids, body size, body mass index and diabetes, cardiovascular traits such as cardiovascular diseases, blood pressure and blood cell measurements, brain traits such as cortical thickness, volume of the diencephalon structures and cerebral white matter, and grey/white matter contrast, bone traits such as density and calcium levels, neurodegenerative diseases, educational attainment, and trauma exposure (Supplementary Table 19). The independent nominally-significant SNPs mapped to multiple genes with two or more methods (*KIAA1614, RP11-46A10.5, RP11-46A10.4, RP11-46A10.8, AL162431.1, STX6, MR1, IER5, RP11-309G3.3, FBRSL1* and *CCNE1*) (Supplementary Table 20).

We used gene-based tests to identify gene-level p-values, however no genes passed genome-wide significance ($P < 2.530e-6$) (Supplementary Figure 18). Gene-set and gene property analyses were performed to determine the functional significance of the GxS analysis. The GxS SNPs were not significantly enriched for any curated gene sets or Gene Ontology terms obtained from MsigDB. SNPs from the GxS interaction analysis were not significantly enriched for gene expression in any tissue types, however skin, muscle and salivary gland were the three most significantly enriched tissues (Supplementary Figure 19).

### Supplementary Figures

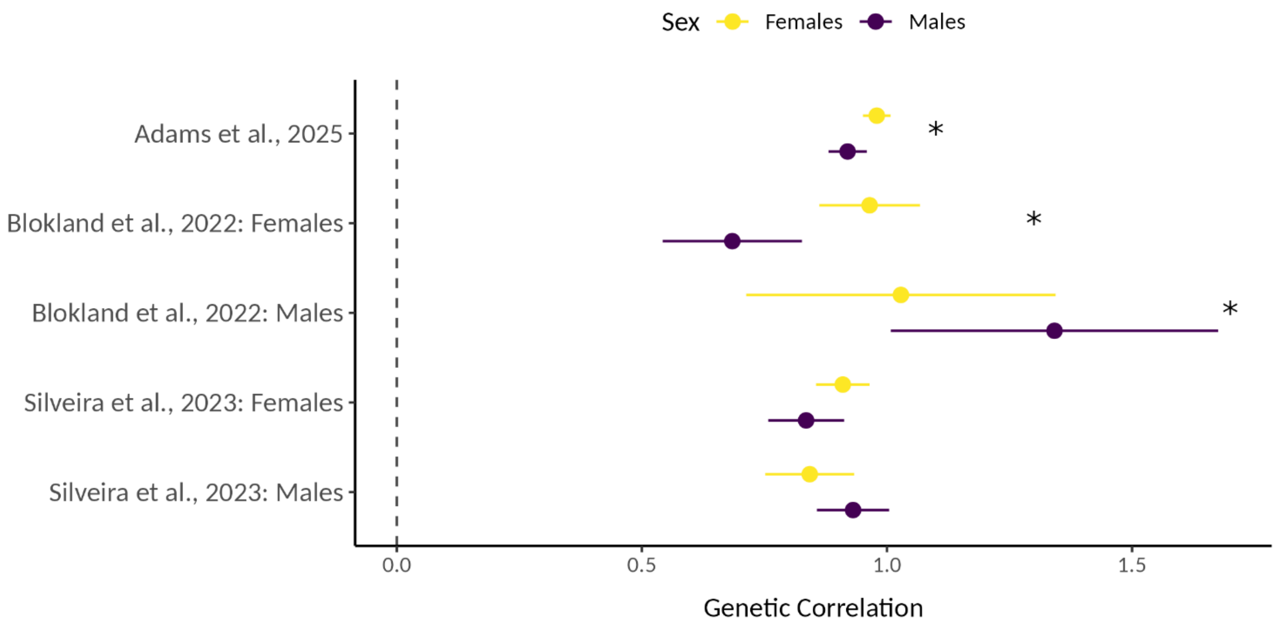


**Supplementary Figure 1. Genetic correlation between our female and male GWAS for depression with the largest previously published GWAS for MDD (Adams *et al.* [3]) and previous sex-stratified GWAS for MDD (Blokland *et al.* [4] and Silveira *et al.* [5]). * = significantly different $r_g$ of our MDD GWAS in females with previously published MDD versus the $r_g$ of our MDD GWAS in males with the same previously published MDD GWAS (after correction of the p-value using the Bengamini-Hochberg method for five tests). The sex-combined GWAS by Adams *et al.* [3] had a significantly higher genetic correlation with our female GWAS, compared to our male GWAS. The female GWAS by Blokland *et al.* [4] had a significantly higher genetic correlation with our female GWAS, compared to our male GWAS. The male GWAS by Blokland *et al.* [4] had a significantly higher genetic correlation with our male GWAS, compared to our female GWAS. Lastly both female and male GWAS by Silveira *et al.* [5] did not show significantly different genetic correlations with our female and male GWAS.**

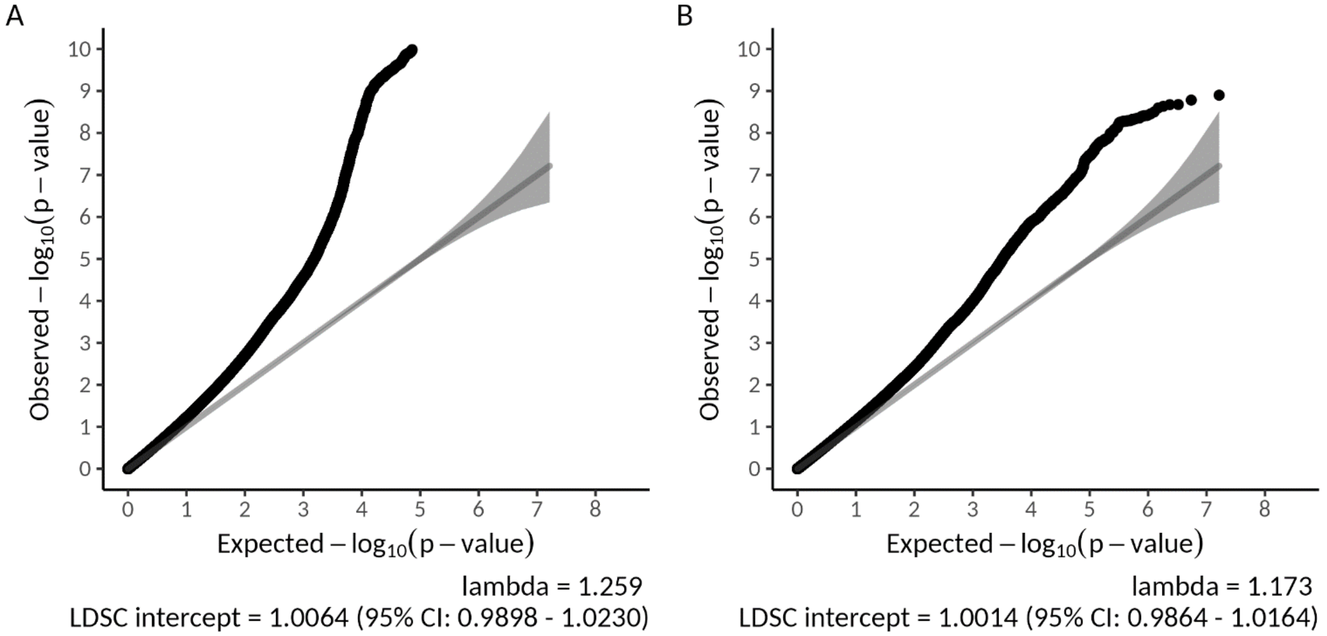


**Supplementary Figure 2. QQ plot, lambda value and Linkage Disequilibrium Score Regression (LDSC) intercept for GWAS meta-analysis in A) females and B) males.**

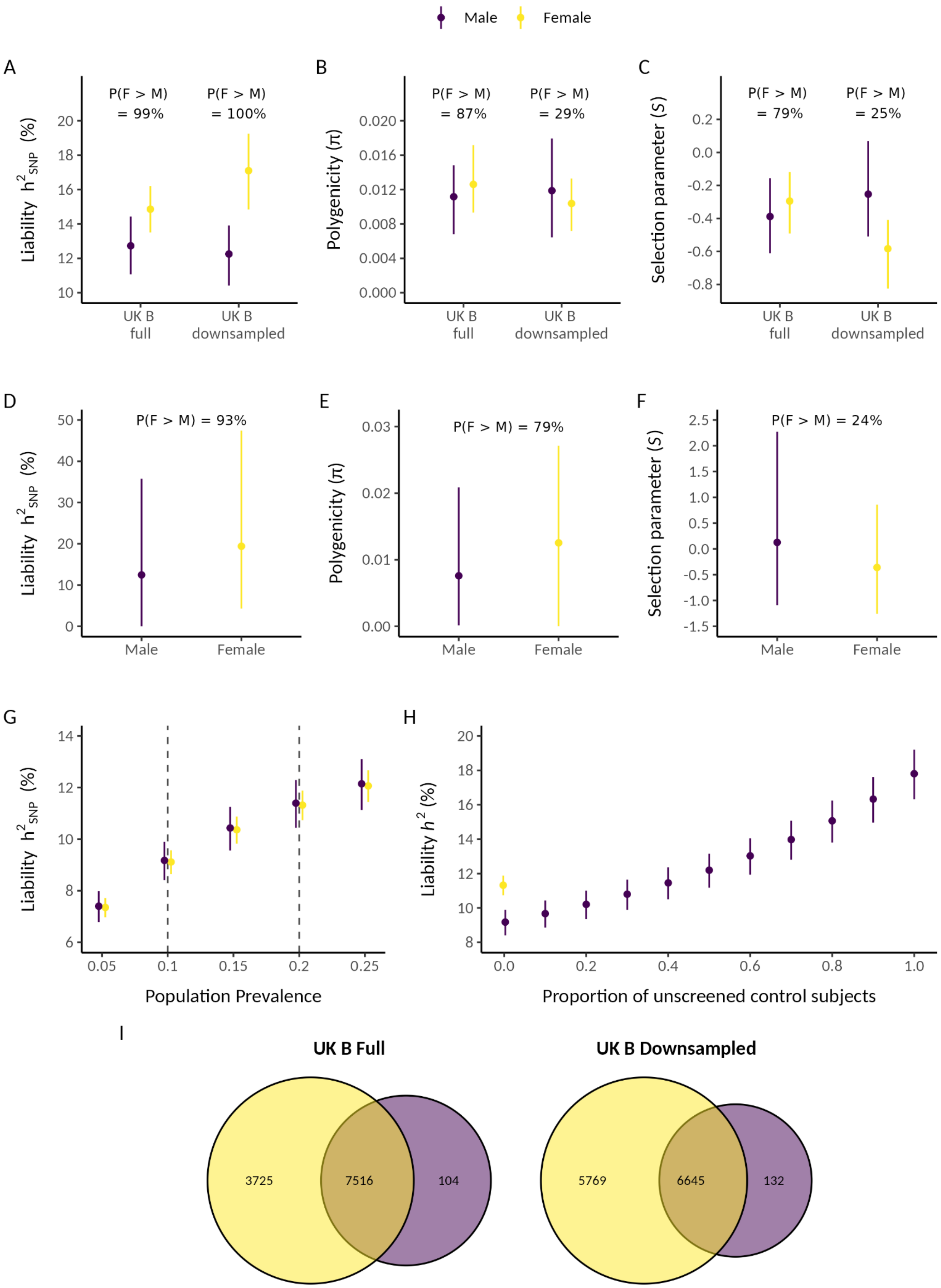
Male
Female
A
P(F > M) = 99%
P(F > M) = 100%
Liability h2SNP (%)
UK B full
UK B downsampled
B
P(F > M) = 87%
P(F > M) = 29%
Polygenicity (π)
UK B full
UK B downsampled
C
P(F > M) = 79%
P(F > M) = 25%
Selection parameter (S)
UK B full
UK B downsampled
D
P(F > M) = 93%
Liability h2SNP (%)
Male
Female
E
P(F > M) = 79%
Polygenicity (π)
Male
Female
F
P(F > M) = 24%
Selection parameter (S)
Male
Female
G
Liability h2SNP (%)
Population Prevalence
H
Liability h2 (%)
Proportion of unscreened control subjects
I
UK B Full
3725
7516
104
UK B Downsampled
5769
6645
132

**Supplementary Figure 3. Sensitivity analyses for comparisons across the two sexes. Sensitivity analyses to test whether our results were influenced by the differential power of our sex-stratified GWAS meta-analysis: Estimates from SBayesS using the full and down-sampled UK Biobank (UK B) cohort for A) Autosomal SNP-based heritability ($h^2_{SNP}$) on the liability scale, B) Polygenicity, and C) Selection parameter. Sensitivity analyses to test whether our results were being driven by across-cohort heterogeneity: Estimates from SBayesS in each of the cohorts separately followed by combination of all Markov chain Monte Carlo (MCMC) samples for D) Autosomal SNP-based heritability ($h^2_{SNP}$) on the liability scale, E) Polygenicity, and F) Selection parameter. G) $h^2_{SNP}$ in females and males using our sex-stratified meta-analysis results across a range of population prevalence's. Vertical dashed lines represent the population prevalence used for males (0.1) and females (0.2) when comparing $h^2_{SNP}$ across sexes. H) $h^2_{SNP}$ on the liability scale for females using a population prevalence of 0.2 and no unscreened controls, and for males using population prevalence of 0.1 – 0.2 corresponding to the proportion of unscreened controls ranging from 0 – 1 (no unscreened controls – all controls are unscreened). I) Venn diagram depicting the number of causal variants explaining 90% of MDD $h^2_{SNP}$ in females only, males only, or both sexes, as identified by MiXeR on the full and down-sampled UK Biobank cohort. Females are in yellow and males in dark purple. As a Bayesian framework was used the points represent the mean posterior value, error bars are the 95% highest posterior density interval and percentages are the posterior probability that female value > male value. (previous page).**

Figure is supplied separately as it is too large to fit in this document.

**Supplementary Figure 4. Pearson correlation between the standardised beta values for each male-female pairwise combination of the six cohorts for the lead independent genome-wide significant SNPs from the largest GWAS meta-analysis of MDD [3]. There are 36 female to male correlations. Meta-analyses were carried out for the six male-female comparisons within the same cohort (plots on the diagonal), and the 30 male-female comparisons across cohorts (plots above and below the diagonals).**

Figure is supplied separately as it is too large to fit in this document.

**Supplementary Figure 5. Pearson correlation between the standardised beta values for each male-male pairwise combination of the six cohorts for the lead independent genome-wide significant SNPs from the largest GWAS meta-analysis of MDD [3]. There are 36 male to male correlations. Meta-analysis was carried out for the 15 unique male-male comparisons across cohorts (plots above the diagonal). All 30 male-male comparisons across cohorts were not included because correlation is not directional, i.e. the 15 plots above the diagonal have the same R as the 15 plots below the diagonal. Male-male comparisons within the same cohort were not included because they have a correlation of 1 (plots on the diagonal).**

Figure is supplied separately as it is too large to fit in this document.

**Supplementary Figure 6. Pearson correlation between the standardised beta values for each female-female pairwise combination of the six cohorts for the lead independent genome-wide significant SNPs from the largest GWAS meta-analysis of MDD [3]. There are 36 female to female correlations. Meta-analysis was carried out for the 15 unique female-female comparisons across cohorts (plots above the diagonal). All 30 female-female comparisons across cohorts were not included because correlation is not directional, i.e. the 15 plots above the diagonal have the same R as the 15 plots below the diagonal. Female-female comparisons within the same cohort were not included because they have a correlation of 1 (plots on the diagonal).**

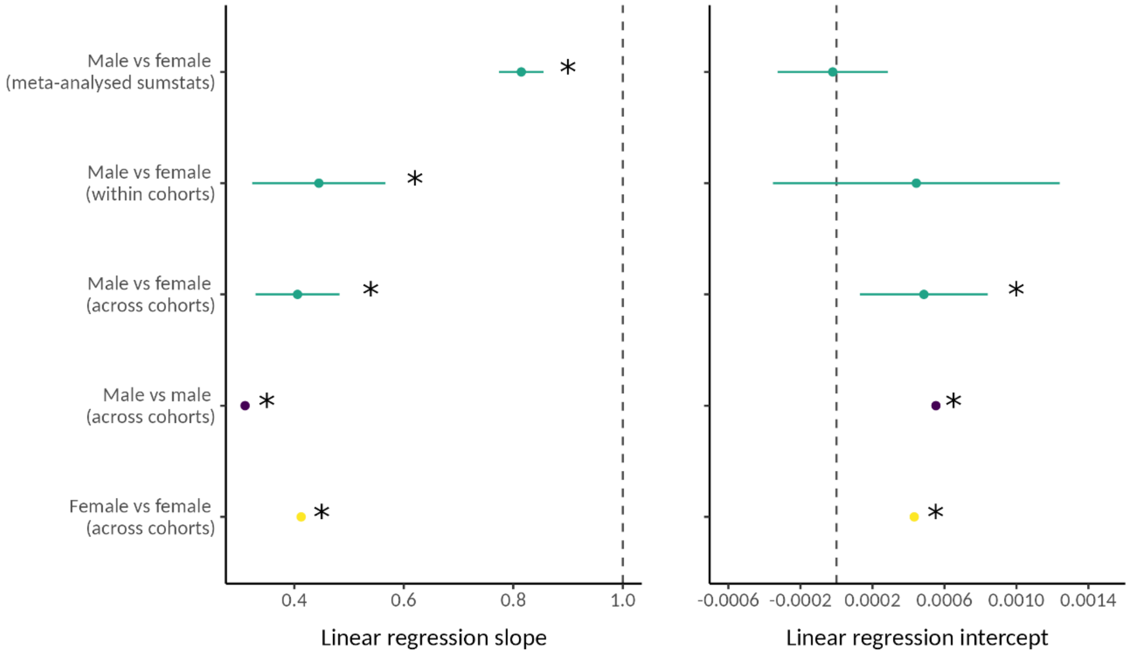


**Supplementary Figure 7. Linear regression A) slope and B) intercept of the MDD effect sizes of SNPs known to be associated with depression for males vs females using our sex-stratified meta-analysis results, and meta-analysis of the slope/intercept estimated in all male-female within cohort and male-female across cohort combinations, as well as the mean slope/intercept from the distribution of the slopes/intercepts from the meta-analyses of all male-male and female-female across cohort comparisons. The points represent the mean while error bars are the 95% confidence interval and stars represent the slope being significantly different to one or the intercept significantly different to zero (after using the Benjamini-Hochberg method to correct the p-value for multiple tests).**

Figure is supplied separately as it is too large to fit in this document.

**Supplementary Figure 8. Linear regression between the standardised beta values for each male-female pairwise combination of the six cohorts for the lead independent genome-wide significant SNPs from the largest GWAS meta-analysis of MDD [3]. There are 36 female to male correlations. Meta-analyses were carried out for the six male-female comparisons within the same cohort (plots on the diagonal), and the 30 male-female comparisons across cohorts (plots above and below the diagonals).**

Figure is supplied separately as it is too large to fit in this document.

**Supplementary Figure 9. Linear regression between the standardised beta values for each male-male pairwise combination of the six cohorts for the lead independent genome-wide significant SNPs from the largest GWAS meta-analysis of MDD [3]. There are 36 male to male comparisons. Unlike correlations, linear regression is directional (A vs B ≠ B vs A). Thus, the 15 plots above the diagonal are not the same as the 15 plots below the diagonal. However, all 30 male-male comparisons cannot be included in the meta-analysis because the linear regressions in both directions are not independent (A vs B is not independent from B vs A). Therefore, a meta-analysis was run for every set of 15 independent linear regressions for male-male comparisons ($2^{15}$ = 32,768 male-male meta-analyses). Male-male comparisons within the same cohort were not included (plots on the diagonal).**

Figure is supplied separately as it is too large to fit in this document.

**Supplementary Figure 10. Linear regression between the standardised beta values for each female-female pairwise combination of the six cohorts for the lead independent genome-wide significant SNPs from the largest GWAS meta-analysis of MDD [3]. There are 36 female to female comparisons. Unlike correlations, linear regression is directional (A vs B ≠ B vs A). Thus, the 15 plots above the diagonal are not the same as the 15 plots below the diagonal. However, all 30 female-female comparisons cannot be included in the meta-analysis because the linear regressions in both directions are not independent (A vs B is not independent from B vs A). Therefore, a meta-analysis was run for every set of 15 independent linear regressions for female-female comparisons ($2^{15}$ = 32,768 female-female meta-analyses). Female-female comparisons within the same cohort were not included (plots on the diagonal).**

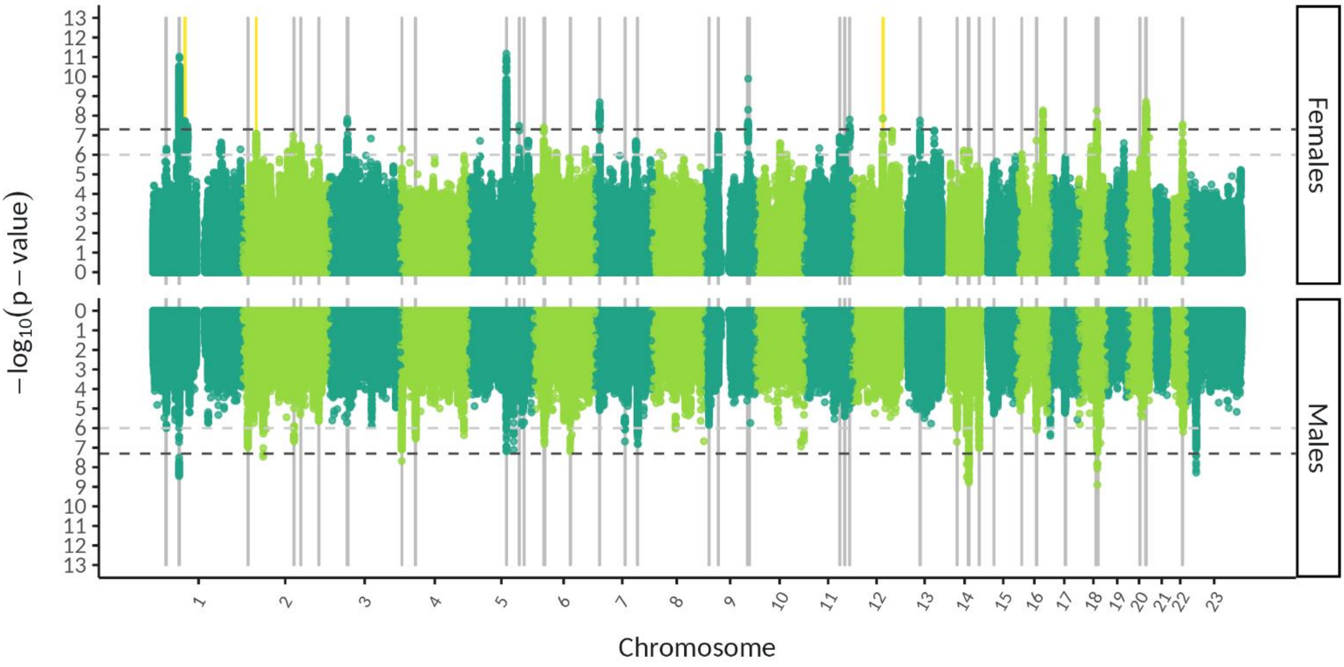


**Supplementary Figure 11. Miami plot of female and male sex-stratified analyses with genomic regions highlighted that were identified in gwas-pw to contain causal risk loci for MDD that are shared between females and males or are sex-specific. Chromosome 23 is the X chromosome (note gwas-pw was used for autosomal SNPs only). Grey bands = evidence from gwas-pw for genomic regions that contain a common causal risk variant for MDD in females and males, yellow bands = evidence from gwas-pw for genomic regions that contain a causal risk variant for MDD only in females.**

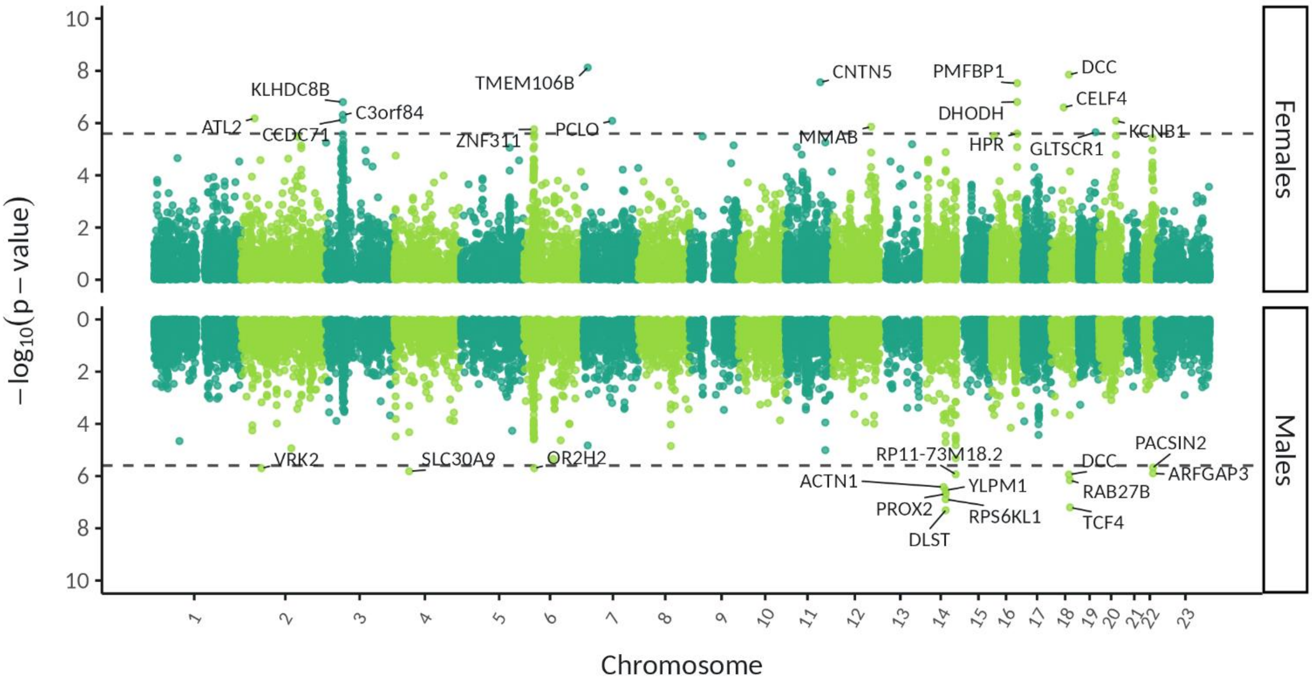


**Supplementary Figure 12. Miami plot of sex-stratified gene-based tests for depression in FUMA. Female and male results are shown on the top and bottom, respectively. Chromosome 23 is the X chromosome. The horizontal dashed line indicates the genome-wide significance P-value of 2.53 x $10^{-6}$ (input SNPs mapped to 19,759 protein coding genes).**

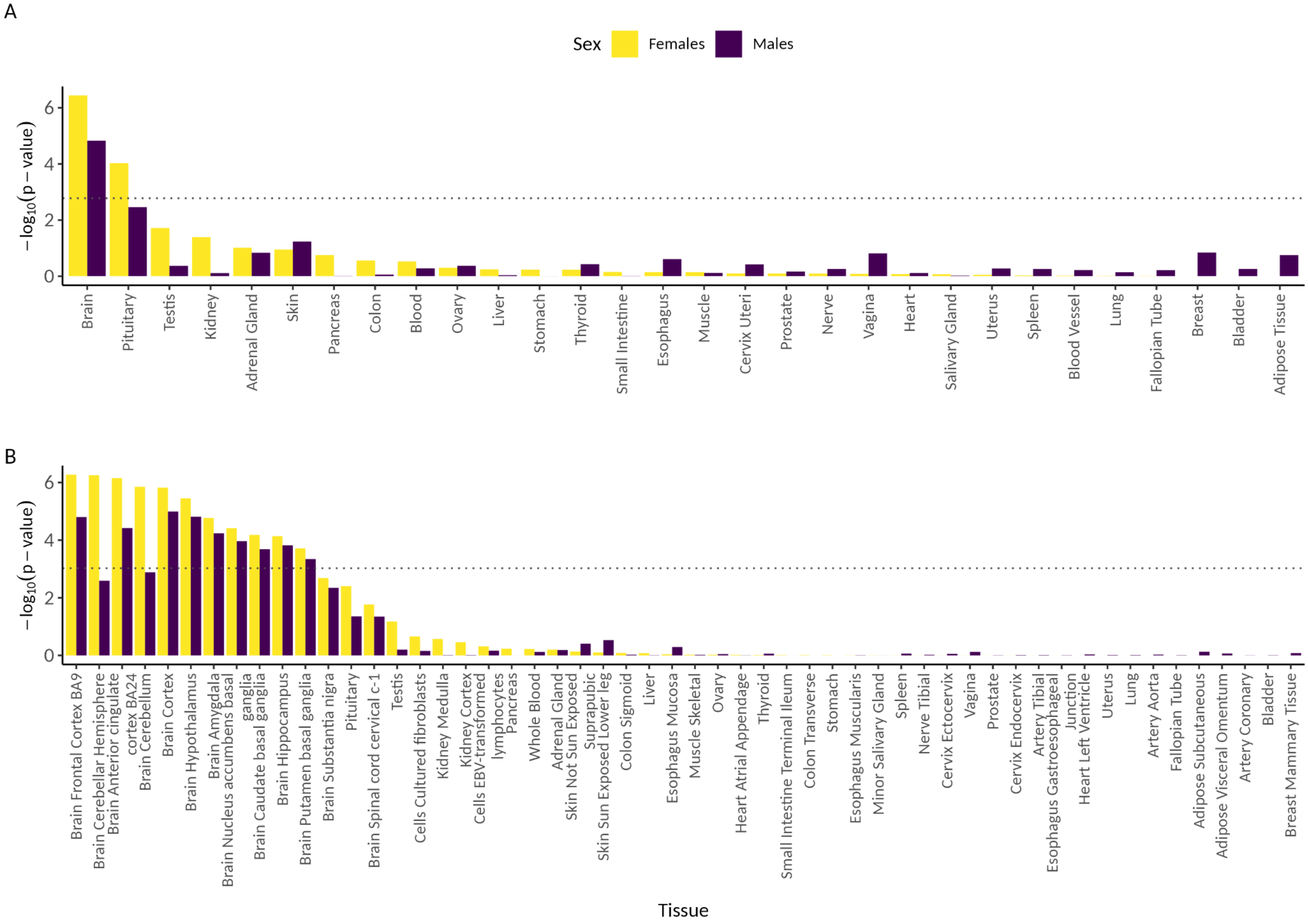
A
Sex
Females
Males
−log10(p − value)
Brain
Pituitary
Testis
Kidney
Adrenal Gland
Skin
Pancreas
Colon
Blood
Ovary
Liver
Stomach
Thyroid
Small Intestine
Esophagus
Muscle
Cervix Uteri
Prostate
Nerve
Vagina
Heart
Salivary Gland
Uterus
Spleen
Blood Vessel
Lung
Fallopian Tube
Breast
Bladder
Adipose Tissue
B
−log10(p − value)
Brain Frontal Cortex BA9
Brain Cerebellar Hemisphere
Brain Anterior cingulate cortex BA24
Brain Cerebellum
Brain Cortex
Brain Hypothalamus
Brain Amygdala
Brain Nucleus accumbens basal ganglia
Brain Caudate basal ganglia
Brain Hippocampus
Brain Putamen basal ganglia
Brain Substantia nigra
Pituitary
Brain Spinal cord cervical c-1
Testis
Cells Cultured fibroblasts
Kidney Medulla
Kidney Cortex
Cells EBV-transformed lymphocytes
Pancreas
Whole Blood
Adrenal Gland
Skin Not Sun Exposed Suprapubic
Skin Sun Exposed Lower leg
Colon Sigmoid
Liver
Esophagus Mucosa
Muscle Skeletal
Ovary
Heart Atrial Appendage
Thyroid
Small Intestine Terminal Ileum
Colon Transverse
Stomach
Esophagus Muscularis
Minor Salivary Gland
Spleen
Nerve Tibial
Cervix Ectocervix
Vagina
Prostate
Cervix Endocervix
Artery Tibial
Esophagus Gastroesophageal Junction
Heart Left Ventricle
Uterus
Lung
Artery Aorta
Fallopian Tube
Adipose Subcutaneous
Adipose Visceral Omentum
Artery Coronary
Bladder
Breast Mammary Tissue
Tissue

**Supplementary Figure 13. Gene-property analysis in FUMA to identify SNPs from the GWAS meta-analysis of MDD in females and males that are significantly enriched for gene expression in A) 30 general tissue types (GTEx v8) and B) 53 tissue types (GTEx v8). The dashed horizontal line represents the Bonferroni corrected significant p-value threshold (0.05/30 and 0.05/53 for A) and B), respectively). Females are in yellow and male in purple. (previous page).**

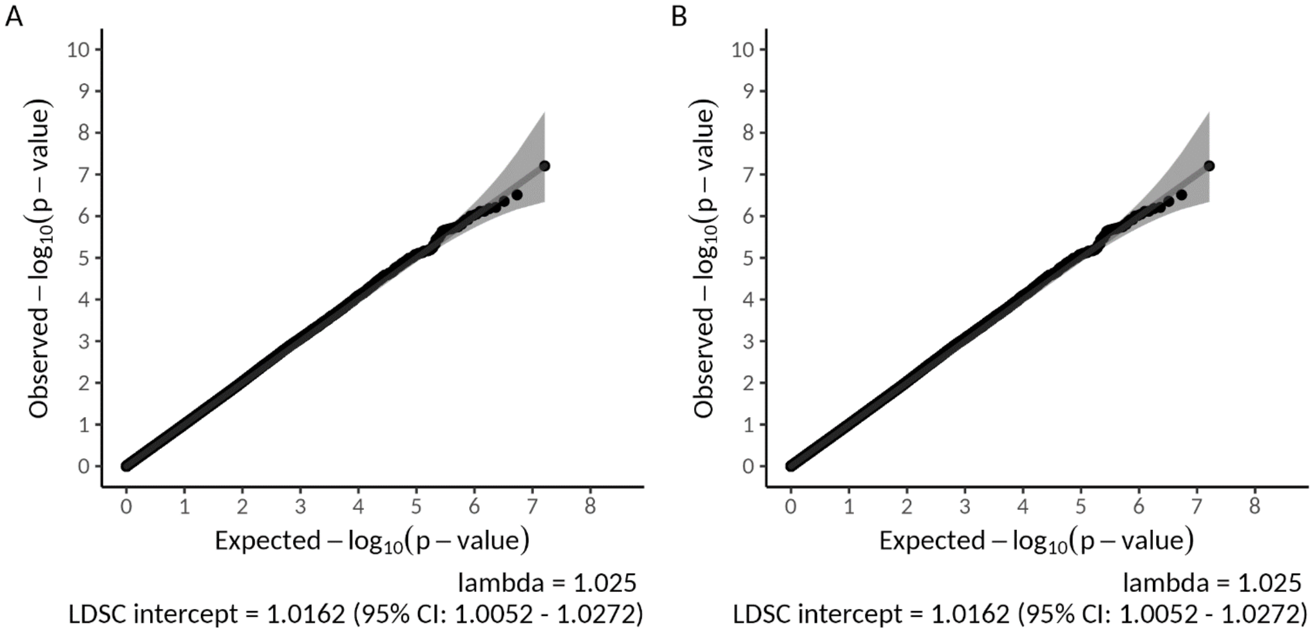


**Supplementary Figure 14. QQ plot, lambda value and Linkage Disequilibrium Score Regression (LDSC) intercept for genome-wide genotype-by-sex interaction meta-analysis using A) full dosage compensation and B) no dosage compensation for SNPs located on the X chromosome non-pseudoautosomal region.**

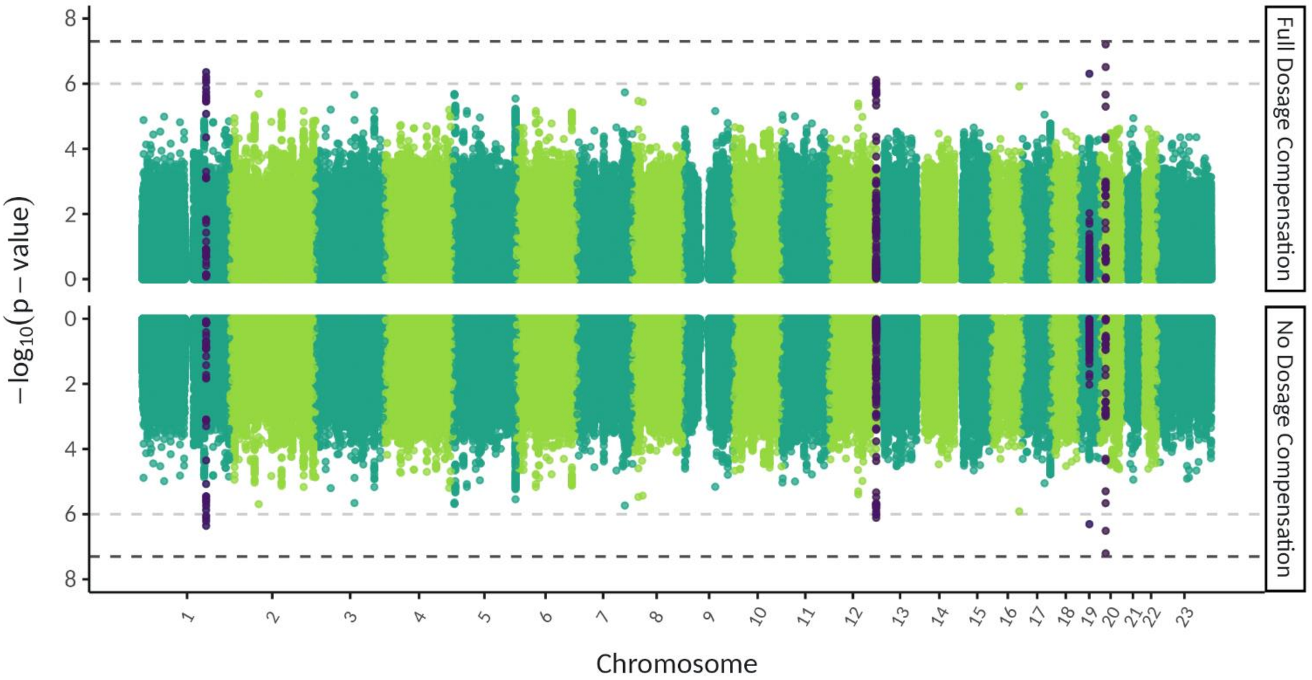


**Supplementary Figure 15. Miami plot of genome-wide genotype-by-sex interaction meta-analysis for depression, with full and no dosage compensation for SNPs on the X chromosome non-pseudoautosomal region shown on the top and bottom, respectively. The –log10 p values for each SNP are shown with positions according to human genome build 37 (GRCh37 assembly). Chromosome 23 is the X chromosome. The darker grey and lighter grey dotted horizontal lines indicate genome-wide significance ($P = 5 \times 10^{-8}$) and nominal significance ($P = 1 \times 10^{-6}$), respectively. SNPs in dark purple indicate the lead independent SNPs that reached nominal significance, and any SNPs in linkage disequilibrium with them.**

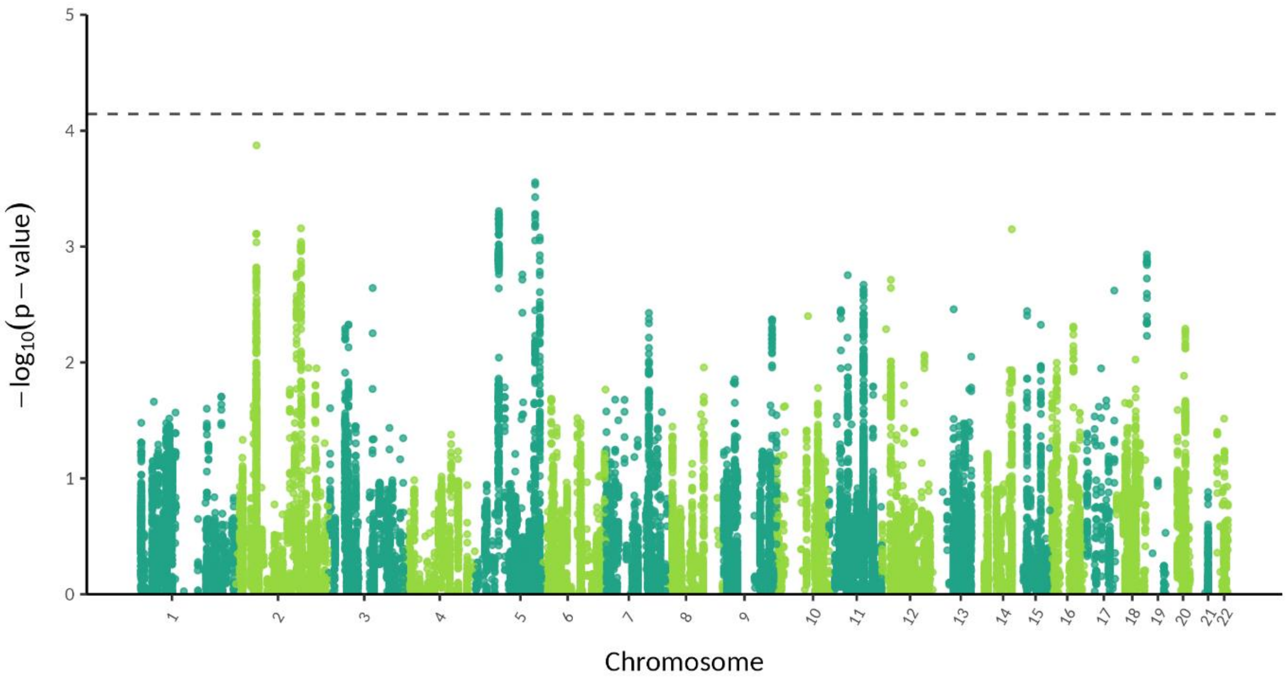


**Supplementary Figure 16. Manhattan plot of genome-wide genotype-by-sex interaction meta-analysis results restricted to the genome-wide significant hits found by Adams *et al.* [3]. Dashed horizontal line is the significance threshold of 0.05/697 (as 697 independent genome-wide significant associations are reported in Adams *et al.* [3]).**

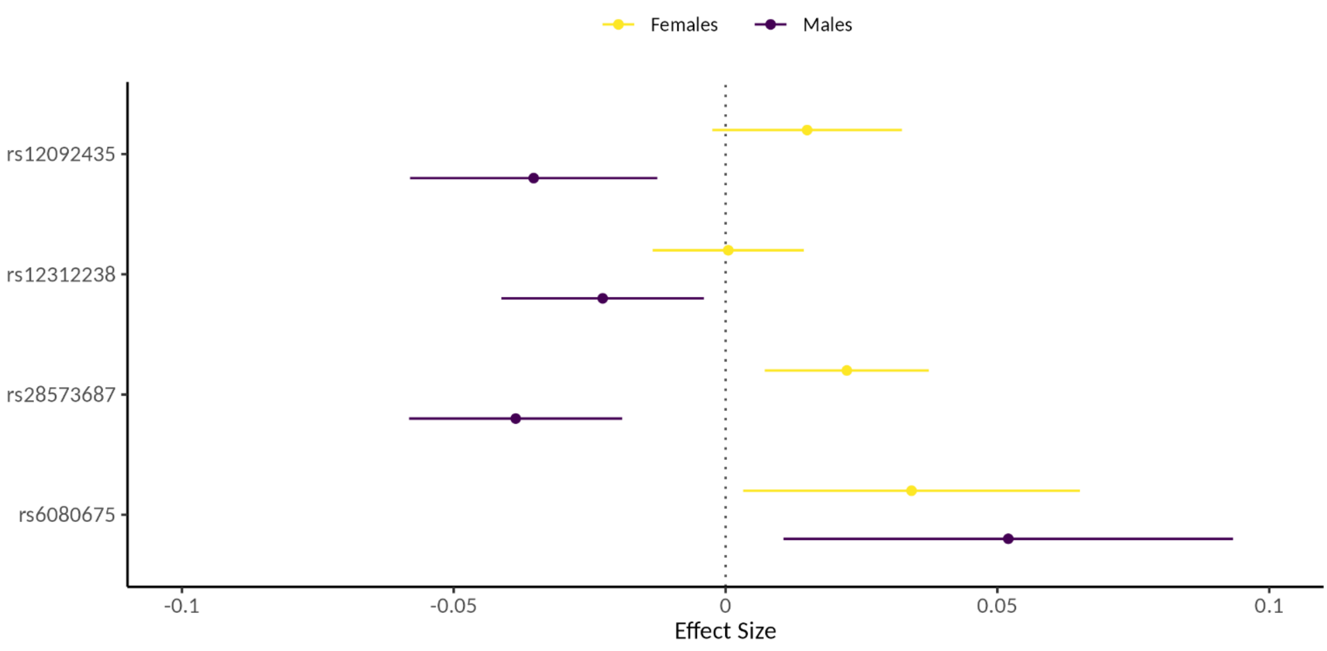


**Supplementary Figure 17. Forest plot showing effect sizes in the sex-stratified GWAS for the independent lead SNPs found to be nominally significant in the GxS analysis. Point and bars are the effect size +/- 95% confidence interval. Females are in yellow and males in dark purple. The dotted vertical grey line is at an effect size of 0, i.e. SNP has no effect on depression.**

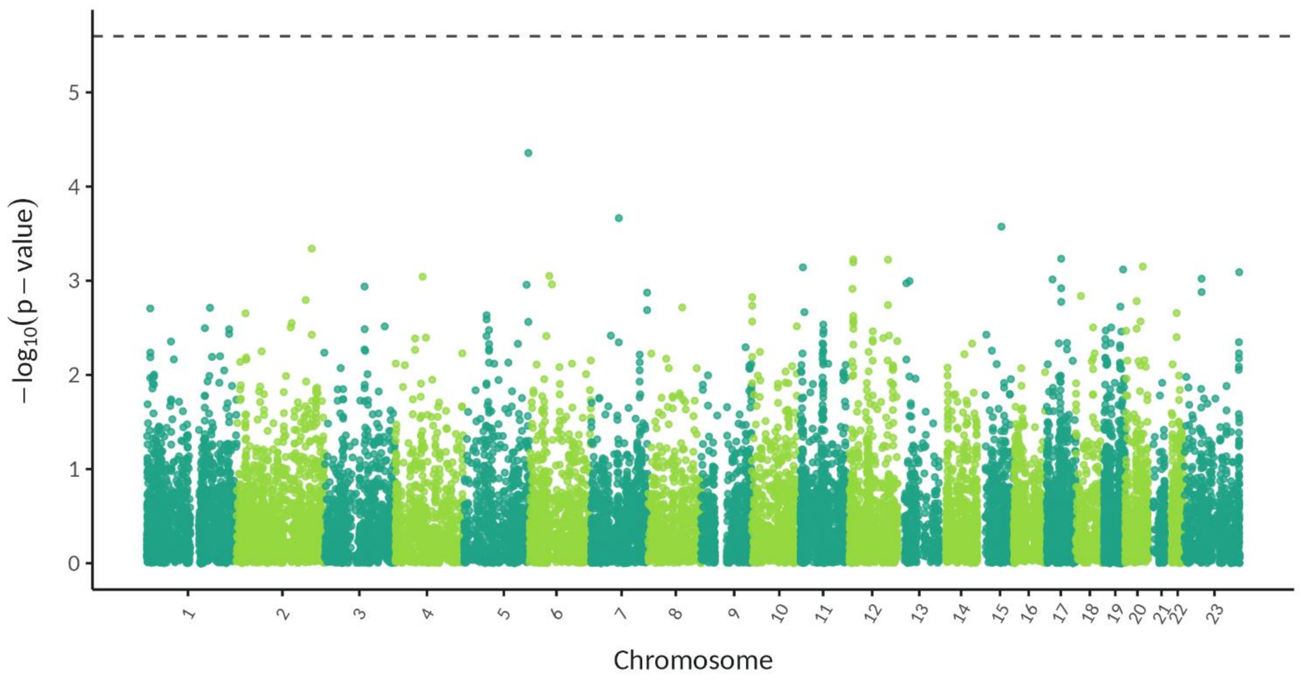


**Supplementary Figure 18. Manhattan plot of genome-wide genotype-by-sex interaction gene-based test for depression in FUMA. Chromosome 23 is the X chromosome. The horizontal dashed line indicates the genome-wide significance P-value of 2.53 x $10^{-6}$ (input SNPs mapped to 19,759 protein coding genes).**

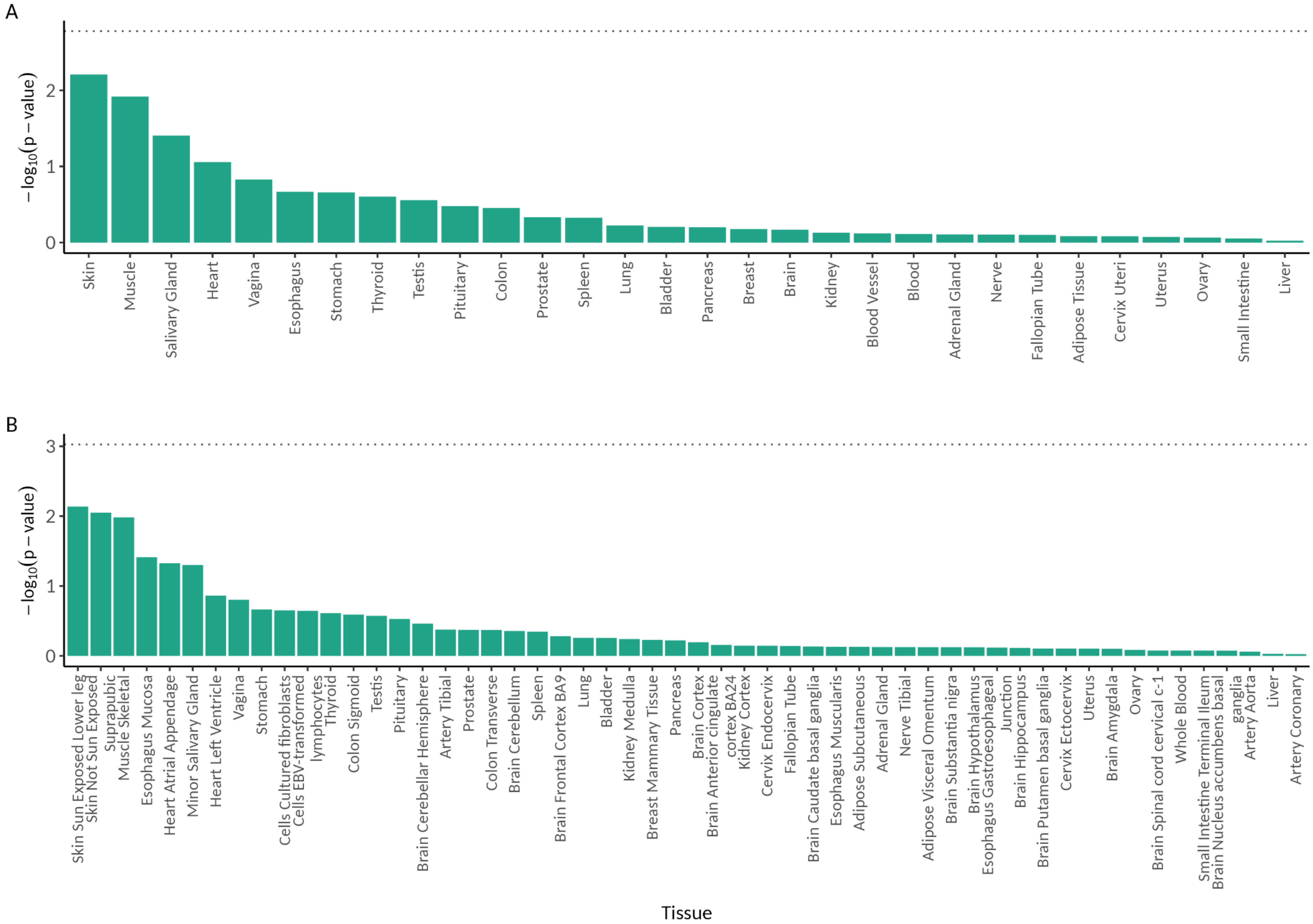
A
−log10(p − value)
0
1
2
Skin
Muscle
Salivary Gland
Heart
Vagina
Esophagus
Stomach
Thyroid
Testis
Pituitary
Colon
Prostate
Spleen
Lung
Bladder
Pancreas
Breast
Brain
Kidney
Blood Vessel
Blood
Adrenal Gland
Nerve
Fallopian Tube
Adipose Tissue
Cervix Uteri
Uterus
Ovary
Small Intestine
Liver
B
−log10(p − value)
0
1
2
3
Skin Sun Exposed Lower leg
Skin Not Sun Exposed Suprapubic
Muscle Skeletal
Esophagus Mucosa
Heart Atrial Appendage
Minor Salivary Gland
Heart Left Ventricle
Vagina
Stomach
Cells Cultured fibroblasts
Cells EBV-transformed lymphocytes
Thyroid
Colon Sigmoid
Testis
Pituitary
Brain Cerebellar Hemisphere
Artery Tibial
Prostate
Colon Transverse
Brain Cerebellum
Spleen
Brain Frontal Cortex BA9
Lung
Bladder
Kidney Medulla
Breast Mammary Tissue
Pancreas
Brain Cortex
Brain Anterior cingulate cortex BA24
Kidney Cortex
Cervix Endocervix
Fallopian Tube
Brain Caudate basal ganglia
Esophagus Muscularis
Adipose Subcutaneous
Adrenal Gland
Nerve Tibial
Adipose Visceral Omentum
Brain Substantia nigra
Brain Hypothalamus
Esophagus Gastroesophageal Junction
Brain Hippocampus
Brain Putamen basal ganglia
Cervix Ectocervix
Uterus
Brain Amygdala
Ovary
Brain Spinal cord cervical c-1
Whole Blood
Small Intestine Terminal Ileum
Brain Nucleus accumbens basal ganglia
Artery Aorta
Liver
Artery Coronary
Tissue

**Supplementary Figure 19. Gene-property analysis in FUMA to identify SNPs from the genome-wide genotype-by-sex interaction analysis that are significantly enriched for gene expression in A) 30 general tissue types (GTEx v8) and B) 53 tissue types (GTEx v8). The dashed horizontal line represents the Bonferroni corrected significant p-value threshold (0.05/30 and 0.05/53 for A) and B), respectively). (previous page).**

**Supplementary Methods A:**

**Cohort Methods**

***AGDS***

***Participants and Phenotype Definition***

Details about the Australian Genetics of Depression Study (AGDS) are published elsewhere [6]. Briefly, over 22,000 participants (approximately 17,000 genotyped) were recruited via Australian government prescription records or through a media campaign. Participants completed online questionnaires, including a core module assessing Major Depressive Disorder (MDD) diagnosis. Here, we used the Composite International Diagnostic Interview short form (CIDI-SF) diagnostic questionnaire [7] to assess MDD DSM-5 criteria. MDD cases were defined as participants in AGDS who met DSM-5 criteria for MDD at some point within their lifetime. That is, during a period of 2+ weeks when the participant's feelings of depression or loss of interest were worst they endorsed at least one of the following two items: 1) felt depressed for most or all of the day for 2+ weeks or 2) had a loss of interest in all or almost all activities every day or almost every day for 2+ weeks. In addition to this, participants reported at least four of the following seven items; 1) weight change or appetite change, 2) hypersomnia or insomnia, 3) fidgety/restless or talking/moving much more slowly, 4) fatigued/less energy, 5) feeling worthless or guilty, 6) difficulty with thinking, concentrating or making decisions and 7) thinking a lot about death.

We used the QSkin Sun and Health Study (QSkin) as a control cohort; a population-based cohort from Queensland, Australia that was invited to participate via a random draw from the electoral roll [8]. Participants completed a lifestyle questionnaire, including a checklist about previous diagnosis, experience or treatment for a range of conditions. MDD controls were defined as QSkin participants who did not report having been diagnosed, experienced or treated for depression nor experienced postnatal or antenatal depression. All protocols and questionnaires for both the AGDS and QSkin cohorts were approved by the QIMR Berghofer Medical Research Institute Human Research Ethics Committee (P2118, P1309 and P2034).

***Genotyping, Quality Control and Imputation***

Participants from both AGDS and QSkin were genotyped using the Illumina Global Screening Array V2. Samples were merged with the 1000 Genomes project samples [9] and genetic principal components (PCs) were calculated in 'smartpca' (build 16000) using a thinned set of single nucleotide polymorphisms (SNPs) (~37,000 SNPs) with the 1000 Genomes individuals used to define the PC axes. Participants without genetic similarity to a European reference group (>6 standard deviations (SD) from Ancestry PCs PC1/PC2 centroid) were excluded. Pre-imputation marker quality control was done using GenomeStudio v2.0 and PLINK 1.9 [10, 11], including removing SNPs with a GenTrain score < 0.6, minor allele frequency < 0.01, SNP call rate < 95%, and deviating from Hardy-Weinberg equilibrium ($p < 1\times10^{-6}$) (restricted to females only for the X chromosome). Imputation was then done using the Haplotype Reference Consortium 1.1 reference panel [12]. To avoid sample overlap in the meta-analysis, all individuals used in Blokland *et al.* [4] were excluded.

##### *Association Analyses*

Directly genotyped autosomal SNPs were filtered and pruned for Linkage Disequilibrium (LD) in PLINK 1.9 [10, 11] using the following flags: --maf 0.01, --geno 0.02, --mind 0.02, --hwe 0.0000000001 and --indep-pairwise 1500 150 0.2. These SNPs were used from European participants only to create a Genetic Relationship Matrix (GRM), followed by a sparse GRM with a threshold of 0.03, using GCTA v1.94.1 [13]. All association analyses were conducted using fastGWA in GCTA v1.94.1 [14]. The first 10 ancestry PCs were used as covariates in both the sex-stratified genome-wide association studies and the genome-wide genotype-by-sex interaction (GxS) analyses.

#### *UK Biobank*

##### *Participants and Phenotype Definition*

The UK Biobank is a cohort study consisting of 488,377 genotyped participants recruited from across the United Kingdom (40 – 69 years at recruitment) [15]. Participants completed a range of questionnaires at various time points, including the Mental health questionnaire (2016) and the Mental well-being questionnaire (2022) both of which included the CIDI-SF. MDD cases were defined as participants that met one or more of the following criteria:

- Met DSM-5 criteria for MDD at some point within their lifetime using the CIDI-SF in the Mental health questionnaire. That is, participants endorsed at least one of the following two items: 1) a depressed mood for 2+ weeks (data field 20446) or 2) loss of interest for 2+ weeks (data field 20441) and they reported feeling down all day long or most of the day (data field 20436) and feeling down every day or almost every day (data field 20439). In addition to this, when thinking about the period in their life, lasting at least two weeks, when their feelings of depression or loss of interest were worst participants reported at least four of the following six items; 1) change in weight (data field 20536), 2) change in sleep (data field 20532), 3) fatigue or loss of energy (data field 20449), 4) feelings of worthlessness (data field 20450), 5) difficulty thinking/concentrating/making decisions (data field 20435), and 6) thoughts about death (data field 20437).

- Met DSM-5 criteria for MDD at some point within their lifetime using the CIDI-SF in the Mental well-being questionnaire. That is, participants endorsed at least one of the following two items: depressed mood for 2+ weeks (data field 29011) or loss of interest for 2+ weeks (data field 29012) and report feeling down all day long or most of the day (data field 29014) and report feeling down every day or almost every day (data field 29015). In addition to this, when thinking about the period in their life, lasting at least two weeks, when their feelings of depression or loss of interest were worst participants reported at least four of the following six items; 1) change in appetite or weight (data field 29020 or 29021), 2) change in sleep (data field 29022), 3) fatigue or loss of energy (data field 29018), 4) feelings of worthlessness or guilt (data field 29027 or 29028), 5) difficulty thinking/concentrating/making decisions (data field 29026), 6) thoughts about death (data field 29029).
- Met the criteria for probable MDD; an answer of 'probable recurrent MDD (severe)' or 'probable recurrent MDD (moderate)' or 'single probable MDD episode' in the data field 20126. This is a derived data field, as explained in Smith *et al.* [16]. Briefly, participants indicated being unenthusiastic/disinterested for ≥ 2 weeks (data fields 4631 and 5375), depressed for ≥ 2 weeks (data fields 4598 and 4609) and having seen a doctor (GP) or psychiatrist for nerves, anxiety, tension or depression (data fields 2090 and 2100).
- Presence of ICD-10 primary and/or secondary codes for depression (data field 41202 and 41204) and self-report of depression (data field 20002). That is, one or more of the ICD-10 codes: F32 (depressive episode), F33 (recurrent depressive disorder), F34 (Persistent mood [affective] disorders) or F39 (Unspecified mood [affective] disorder) in data field 41202 or 41204, and code 1286 in data field 20002.

MDD controls were defined as participants meeting all of the following criteria:

- Do not meet the DSM-5 criteria for MDD at some point within their lifetime using the CIDI-SF in the Mental health questionnaire.
- Do not meet the DSM-5 criteria for MDD at some point within their lifetime using the CIDI-SF in the Mental-wellbeing questionnaire.
- Do not meet the criteria for probable MDD; an answer of 'no bipolar or depression' in the data field 20126.
- No ICD-10 primary and/or secondary codes for depression reported. That is, no report of F32, F33, F34 or F39 in data fields 41202 and 41204.
- No self-report of depression. That is no code 1286 in data field 20002.

However, if participants did not answer the CIDI-SF in the mental health and/or mental well-being questionnaires they were still defined as controls if the remaining three criteria were met, and if participants did not answer the questions needed to determine probable MDD they were still defined as controls if the remaining three criteria were met.

##### *Genotyping, Quality Control and Imputation*

We used the genotype data after quality control and imputation as provided by the UK Biobank [15]. Individuals who withdrew consent and not defined as genetic white-British ancestry by the UK Biobank were excluded.

##### *Association Analyses*

Directly genotyped autosomal SNPs were filtered and pruned for Linkage Disequilibrium (LD) in PLINK 1.9 [10, 11] using the following flags --maf 0.01, --geno 0.02, --mind 0.1, --hwe 0.0000000001 and --indep-pairwise 1500 150 0.2. These SNPs were used from participants with white-British ancestry only to create a Genetic Relationship Matrix (GRM), followed by a sparse GRM with a threshold of 0.05, using GCTA v1.94.1 [13]. All association analyses were conducted using fastGWA in GCTA v1.94.1 [14]. The first 10 ancestry PCs were used as covariates in both the sex-stratified genome-wide association studies and the GxS analyses.

#### *All Of Us*

##### *Participants and Phenotype Definition*

The All Of Us Research Program is an ongoing cohort study consisting of over 400,000 participants at present recruited from across the United States (18 years and older at enrolment) [17]. Participants provided survey responses on lifestyle, demographics, and health history, linked Electronic Health Records (EHR), physical measurements, and biospecimens. Whole-genome sequencing data are available for a subset of participants (~245,000 at present), with ongoing data releases expanding coverage [18].

Participants that completed the Personal and Family Health History survey self-reported whether they had ever been diagnosed with a mental health or substance use condition (Concept Code: 43529217). MDD cases were defined as participants that self-reported they had personally been diagnosed with depression (Concept Code: 1384656), or the presence of an EHR code for 'major depressive disorder' (SNOMED:370143000). MDD controls were defined as the absence of an electronic health record for depression and no self-report of depression.

#### *Genotyping, Quality Control and Imputation*

We used the All of Us short read whole genome SNP & Indel smaller callset (ACAF v7.1). The All Of Us ancestry prediction was used to exclude participants of non-European genetic ancestry. Participants with genotype missingness > 0.05, discordant self-reported sex and genetic sex (using genomic_metrics.tsv provided by All Of Us) and with at least one outlier metric (using flagged_samples.tsv provided by All Of Us) were also excluded. Marker quality control included removing SNPs with minor allele frequency < 0.01 and missingness > 0.05. On the autosomes only, SNPs deviating from Hardy-Weinberg equilibrium ($p < 1\times10^{-10}$) were also removed.

#### *Association Analyses*

Autosomal SNPs were filtered and pruned for Linkage Disequilibrium (LD) in PLINK 2 [10, 11] using the following flags --maf 0.01, --geno 0.02, --mind 0.05, --hwe 0.0000000001 and --indep-pairwise 1500 150 0.2. These SNPs were used from participants with European ancestry only to create a Genetic Relationship Matrix (GRM), followed by a sparse GRM with a threshold of 0.05, using GCTA v1.94.1 [13]. All association analyses were conducted using fastGWA in GCTA v1.94.1 [14]. The first 10 ancestry PCs were used as quantitative covariates and the sequencing site as a discrete covariate in both the sex-stratified genome-wide association studies and the GxS analyses.

#### ***BIONIC***

##### ***Participants and Phenotype Definition***

The BIObanks Netherlands Internet Collaboration (BIONIC) is a consortium of 16 Dutch studies and biobanks aiming to characterize the genetics of depression in the Netherlands. Cohorts include a mix of clinical and general populations who were approached for harmonious DSM-5 MDD data collection through the Lifetime Depression Assessment Survey (LIDAS) [19]. These data, together with previously collected DSM-5 MDD data and genotype data, were aggregated at a central location and harmonized (identical phenotype definition and genotype QC and imputation) for analysis [20]. Cases were defined as individuals who met DSM-5 criteria for MDD at some point within their lifetime using the LIDAS, CIDI or Mini-International Neuropsychiatric Interview (MINI) questionnaires. That is, ever had a period of at least two weeks where an individual reported feeling down or anhedonia every day or almost every day and report four or more of the following symptoms when thinking about the period in their life, lasting at least two weeks, when their feelings of depression or loss of interest were worst: change in weight, change in sleep, fatigue or loss of energy, feelings of guilt or worthlessness, difficulty thinking/concentrating/making decisions, thoughts about death. Controls were defined as individuals who did not meet these criteria or who had a low symptom score (<10) on the Center for Epidemiologic Studies Depression Scale (CES-D), Adult Self-report - The Achenbach System of Empirically Based Assessment (ASR-ASEBA), Beck's Depression Inventory (BDI), or Hospital Anxiety and Depression Scale (HADS). Controls were screened for a diagnostic or treatment history of psychopathology and antidepressant use when such information was available.

##### ***Genotyping, Quality Control and Imputation***

BIONIC participants were genotyped using Affymetrix 6, Axiom Finngen, Axiom-NL, Illumina CytoSNP, Global Screening Array, Human Core Exome, and Omnichip. Pre-imputation sample and marker quality control was done using PLINK (v1.9) [10, 11] and KING (v2.2.6) [21], and included discrepancy between reported and biological sex (PLINK FchrX-coefficient < 0.8 for males and FchrX > 0.2 for females), excess heterozygosity (Fautosomes > 0.10 or < -0.10), insufficient sample call rate (< 0.90) and call rate by chromosome (< 0.80), or incorrect identity-by-descent sharing between relatives. SNPs were excluded based on Hardy Weinberg Equilibrium ($p < 1x10^{-4}$) and SNP call rate (< 0.95), as well as Mendelian error rates above 1%. Palindromic SNPs were excluded with minor allele frequency > 0.30. SNPs were aligned to the Haplotype Reference Consortium (HRC) panel (v1.1) [12] and SNPs with an allele frequency difference > 0.10 with the reference data were also excluded. Imputation was then done using the HRC reference panel (v1.1). Principal component analysis was conducted in PLINK (v1.9) on the imputed SNP data based on the three superpopulations (African, Asian, European) from the 1000 Genomes Project reference panel (phase 3v5) [9]. The BIONIC genotype data were SNP and LD pruned and projected onto the 1000 Genomes Project PCA space to compute principal components (PCs). Participants without genetic similarity to a European reference group (>4 standard deviations (SD) from the first six ancestry PC centroids) were excluded. To avoid sample overlap in the meta-analysis, all individuals included in Blokland *et al.* [4] were excluded.

##### *Association Analyses*

Directly genotyped autosomal SNPs were filtered and pruned for Linkage Disequilibrium (LD) in PLINK 1.9 [10, 11] using the following flags --maf 0.01, --geno 0.01, --mind 0.1, and --indep-pairwise 50 5 2. These SNPs were used from participants with European ancestry only to create a Genetic Relationship Matrix (GRM), followed by a sparse GRM with a threshold of 0.05, using GCTA v1.94.1 [13]. All association analyses were conducted using fastGWA in GCTA v1.94.1 [14]. The first 10 ancestry PCs and age were used as quantitative covariates in both the sex-stratified genome-wide association studies and the genome-wide genotype-by-sex interaction (GxS) analyses.

#### *GLAD+*

##### *Participants and Phenotype Definition*

The GLAD+ Study combines two United Kingdom (UK) cohorts: the Genetic Links to Anxiety and Depression (GLAD) Study (www.gladstudy.org.uk) [22], and the National Institute for Health and Care Research (NIHR) BioResource COVID-19 Psychiatry and Neurological Genetics (COPING) Study. The ongoing GLAD Study recruits participants with depression and/or anxiety. Participants provide demographic, environmental, and genetic data and consent to medical record linkage and recontact. The GLAD study comprises >64,000 consented participants, with >50,000 having completed online surveys, and >35,000 having given saliva samples. During the COVID-19 pandemic, the GLAD Study research team recontacted GLAD participants and healthy volunteers from other NIHR BioResource (https://bioresource.nihr.ac.uk/) studies to conduct the COPING study, including >20,000 participants with psychiatric disorders and >11,000 healthy volunteers, two-thirds of whom have been genotyped. Participants completed online questionnaires, including a core module assessing Major Depressive Disorder (MDD) diagnosis. Here, we used the Composite International Diagnostic Interview short form (CIDI-SF) diagnostic questionnaire to assess MDD DSM-5 criteria. MDD cases were defined as participants in GLAD who met DSM-5 criteria for MDD at some point within their lifetime. MDD controls were defined as participants who did not meet DSM-5 criteria for MDD at any point within their lifetime. Most MDD cases (78%) were originally enrolled in the GLAD Study, while all controls were from the COPING study.

#### *Genotyping, Quality Control and Imputation*

Participants from GLAD+ were genotyped by ThermoFisher on the UK Biobank Axiom Array v1 and v2 across numerous genotyping batches. Ancestry was determined using GenoPred (https://opain.github.io/GenoPred/index.html), by projecting GLAD+ individuals on genomic principal components from the 1000 Genomes reference data, and assigning individuals a genetic ancestry if they lay < 3 SD from the mean of individuals from that ancestry superpopulation in 1000 Genomes. Quality control was conducted, excluding variants with MAF < 0.01, call rate < 0.95, or which were deviant from Hardy-Weinberg equilibrium ($p < 1 \times 10^{-10}$). Individuals were excluded if they had withdrawn from the study following genotyping, if they were a duplicate of a higher-quality sample (not including known identical twins), if they were known to be mislabelled, if their genotypic sex (males Fx > 0.8, females Fx < 0.5) did not match their sex assigned at birth, if they were outliers on genome-wide heterozygosity (absolute(Fhat) > 0.2), or if they had an excess of relatives (average pi-hat > 3 SD from the mean). Following quality control, 33,635 individuals and 484,182 variants were available for imputation. Imputation was carried out to TopMED Freeze 8, using the dedicated imputation server ( https://imputation.biodatacatalyst.nhlbi.nih.gov/#!). Following imputation, data was further restricted to data with MAF >= 0.01 and R2 >= 0.3, leaving 15,009,228 variants for analysis. Only individuals assigned European genetic ancestry were used and to avoid sample overlap in the meta-analysis, all Individuals also included in the UK Biobank were excluded (n=1,633).

#### *Association Analyses*

Directly genotyped autosomal SNPs were filtered and pruned for Linkage Disequilibrium (LD) in PLINK 1.9 [10, 11] using the following flags --maf 0.01, --geno 0.02, --mind 0.02, --hwe 0.0000000001 and --indep-pairwise 1500 150 0.2. These SNPs were used from participants with European ancestry only to create a Genetic Relationship Matrix (GRM), followed by a sparse GRM with a threshold of 0.05, using GCTA v1.94.1 [13]. All association analyses were conducted using fastGWA in GCTA v1.94.1 [14]. The first 10 ancestry PCs were used as quantitative covariates and the genotyping batch as a discrete covariate in both the sex-stratified genome-wide association studies and the genome-wide genotype-by-sex interaction (GxS) analyses.

### *Generation Scotland (Replication Cohort)*

#### *Participants and Phenotype Definition*

Generation Scotland is a cohort study of 7,000 families recruited from the general population of Scotland [23]. All clinical participants were screened for a history of emotional and psychiatric disorders using the structured clinical interview for DSM-IV disorders (SCID) [24, 25]; 21.7% screened positive and were invited to continue the interview that focused on mood disorders; of these, 88% completed the interview (19.0% of participants). A subset participated in an online follow-up that included a CIDI (Composite International Diagnostic Interview) [26]. Here, we used the SCID or CIDI to assess MDD DSM-5 criteria. MDD cases were defined as participants in Generation Scotland who met DSM-5 criteria for MDD at some point within their lifetime using the SCID-5 or CIDI. MDD controls were defined as participants who did not meet DSM-5 criteria for MDD at any point within their lifetime. A total of 2,441 cases and 3,321 controls in females, and 938 cases and 2,491 controls in males were used for the replication analysis. Ethical approval for the original data collection was obtained from the Tayside Committee on Medical Research Ethics A (ref 05/S1401/89). Generation Scotland is currently approved as a Research Tissue Bank by the East of Scotland Research Ethics Service (ref 20/ES/0021).

#### *Genotyping, Quality Control and Imputation*

Participants from Generation Scotland were genotyped using the Illumina OmniExpress array, and imputed using the Haplotype Research Consortium (HRC) dataset. Genotyping quality control was performed using the following procedures: individuals with a call rate less than 98% were removed, as were SNPs with a call rate less than 98% or Hardy-Weinberg equilibrium *p* value less than $1 \times 10^{-6}$ (autosomes) or $1 \times 10^{-5}$ (X chromosome). Mendelian errors, determined using relationships recorded in the pedigree, were removed by setting the individual-level genotypes at erroneous SNPs to missing. Ancestry outliers who were more than six standard deviations away from the mean, in a principal component analysis of Generation Scotland merged with 1,092 individuals from the 1000 Genomes Project, were excluded. Further details of methods are described in Nagy *et al.* [27]. A total of 20,032 individuals (8,227 male participants and 11,805 female participants) passed all quality control thresholds. The number of genotyped autosomal SNPs that passed all quality control parameters was 604,858. To avoid sample overlap, all individuals also included in the UK Biobank were excluded.

#### *Association Analyses*

Directly genotyped autosomal SNPs were filtered and pruned for Linkage Disequilibrium (LD) in PLINK 2 [10, 11] using the following flags --maf 0.01, --geno 0.02, --mind 0.02, --hwe 0.0000000001 and --indep-pairwise 1500 150 0.2. These SNPs were used from participants with European ancestry only to create a Genetic Relationship Matrix (GRM), followed by a sparse GRM with a threshold of 0.05, using GCTA v1.94.1 [13]. Sex-stratified association analyses were run using fastGWA in GCTA v1.94.1 [14] using a mixed linear model for a binary outcome (--fastGWA-mlm-binary) with a sparse genetic relationship matrix and the first four ancestry principal components as covariates. The beta-values were compared from our meta-analysis to the replication analysis for the lead independent genome-wide significant SNPs, looking for concordant signs.

**Supplementary Methods B:**

**Sensitivity Analyses for the Genetic Architecture of Depression in Females and Males**

***LDSC***

We also estimated $h^2_{SNP}$ in Linkage Disequilibrium Score Regression (LDSC) (v1.0.1) [28]. Our sex-stratified meta-analysis summary statistics, European LD scores computed from 1000 Genomes and a population prevalence of 0.2 and 0.1 in females and males, respectively, were used. The Z-score (Formula 1), followed by the p-value calculated from a standard normal distribution using a two-tailed test was used to determine whether $h^2_{SNP}$ was significantly different across sexes.

$$Z = \frac{h^2(female) - h^2(male)}{\sqrt{(SE^2(female) + SE^2(male))}}$$

**Formula 1**

***Differential Power***

As our female GWAS has a 1.65-fold larger effective sample size compared to that in males ($n_{eff\ (Females)}$ = 287,082; $n_{eff\ (Males)}$ = 173,943) we also tested whether our results were influenced by this power difference. We ran SBayesS, as described in the main text methods, on the full UK Biobank sample ($n_{Females}$ = 46,194 cases and 53,211 controls, $n_{Males}$ = 22,608 cases and 56,516 controls) and after down-sampling (n = 22,608 cases and 53,211 in both females and males).

***Male Under-diagnosis***

For common disorders, such as MDD, $h^2_{SNP}$ on the liability scale can be underestimated when the controls are not screened, i.e. the controls are contaminated with cases [1]. Some studies report that male MDD is under diagnosed [2]. Therefore, we estimated $h^2_{SNP}$ on the liability scale for males using the equation from Peyrot *et al.* [1] with the proportion of unscreened controls ranging from 0 – 1 (i.e. all controls screened – no screening of controls) and the corresponding increase in population prevalence from 0.1 – 0.2 (Supplementary Table 10).

***Across-cohort Heterogeneity***

As a sensitivity analysis, autosomal SNP-based heritability ($h^2_{SNP}$), polygenicity ($\pi$) and the selection parameter ($S$) were estimated using SBayesS in males and females for each of the six cohorts separately (AGDS, All Of Us, Bionic, GLAD+, UK Biobank and Blokland *et al.* [4]). Due to convergence issues, problematic SNPs identified by the quality control tool DENTIST v1.3 [29] were excluded when running SBayesS for the female summary statistics from Blokland *et al.* [4], priors for $h^2_{SNP}$ and π were set at 0.06 and 0.01, respectively, for the male summary statistics from GLAD+, and SNPs with a sample size in the lowest 30% quantile were removed in the male summary statistics from Blokland *et al.* [4] (SNPs with N < 15,871 were removed. Maximum N = 27,904). All Markov chain Monte Carlo (MCMC) samples were combined across the six studies in females and males separately and the frequency of MCMC samples in which female value > male value was calculated to determine the posterior probability.

**Supplementary Methods C:**

**Male vs Female Linear Regression**

As well as using Pearson's correlation to investigate whether the MDD effect sizes (betas) for SNPs known to be associated with sex-combined MDD are different across the sexes, we also explored the slope and intercept values from linear regressions. For independent, genome-wide significant SNPs associated with MDD [3], we ran a linear regression between the standardised effect size estimates (beta values) of our male versus female meta-analysis summary statistics, and between each pairwise combination of the two sexes by six studies. Meta-analyses were done for the slope and intercept of each linear regression for the six male-female comparisons within studies and the 30 male-female comparisons across studies. Unlike correlations, linear regression is directional (A vs B ≠ B vs A). Thus, for within sex comparisons both linear regression results should be included in the meta-analysis (30 linear regressions for same sex comparisons across the six studies). However, linear regressions in both directions are not independent (A vs B is not independent from B vs A). Therefore, a meta-analysis was run for every set of 15 independent linear regressions for female-female and male-male comparisons ($2^{15}$ = 32,768 female-female meta-analyses and 32,768 male-male meta-analyses) (Supplementary Figures 8 – 10). To determine whether the observed male-female across-cohort linear regression slope/intercept differs significantly from the distribution of male-male or female-female slopes/intercepts the Z-score was calculated (Formula 2), where $\hat{\beta}_{MF}$ = Estimated slope or intercept for the male vs female linear regression, $\bar{\beta}_{reference}$ = Mean slope or intercept from the reference distribution (male vs male or female vs female meta-analyses), $SD(\beta_{reference})$ = Standard deviation of the slopes or intercepts in the reference distribution. The P-value was calculated from a standard normal distribution using a two-tailed test and adjusted for two comparisons using the Benjamini-Hochberg method.

$$Z = \frac{\hat{\beta}_{MF} - \bar{\beta}_{reference}}{SD(\beta_{reference})}$$

**Formula 2**

**Supplementary Methods D:**

**Z-Score Method for Sex-Specific Genetic Correlations**

To determine whether $r_g$ between MDD and each trait is significantly different across sexes we used the jack-knife method, as described in the main text. As a sensitivity analysis we also used the Z-score method as it is commonly used. However, the Z-score assumes that the two genetic correlation estimates are independent, which is not appropriate here as the same second trait is used in both the genetic correlations being compared (e.g. MDD in females versus ADHD compared to MDD in males versus ADHD). The Z-score was calculated (Formula 3) and the corresponding P-value obtained using a standard normal distribution with a two-tailed test followed by the Benjamini Hochberg correction for multiple tests.

$$Z = \frac{r_g(female) - r_g(male)}{\sqrt{\left(SE^2(female) + SE^2(male)\right)}}$$

**Formula 3**
