## Supplementary figures and images for "Sex-stratified genome-wide association meta-analysis of Major Depressive Disorder"

### Supplementary Figure 4

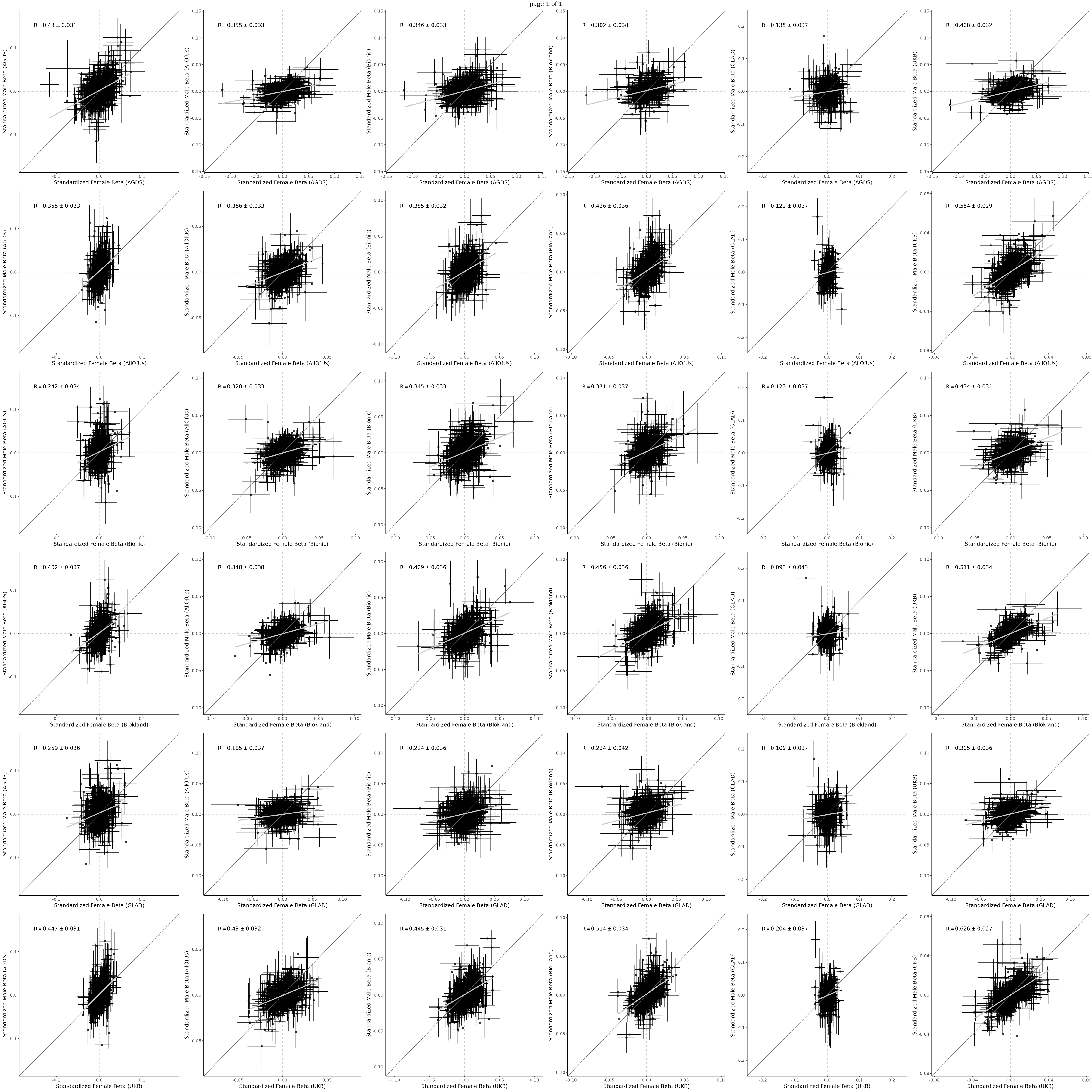

### Supplementary Figure 5

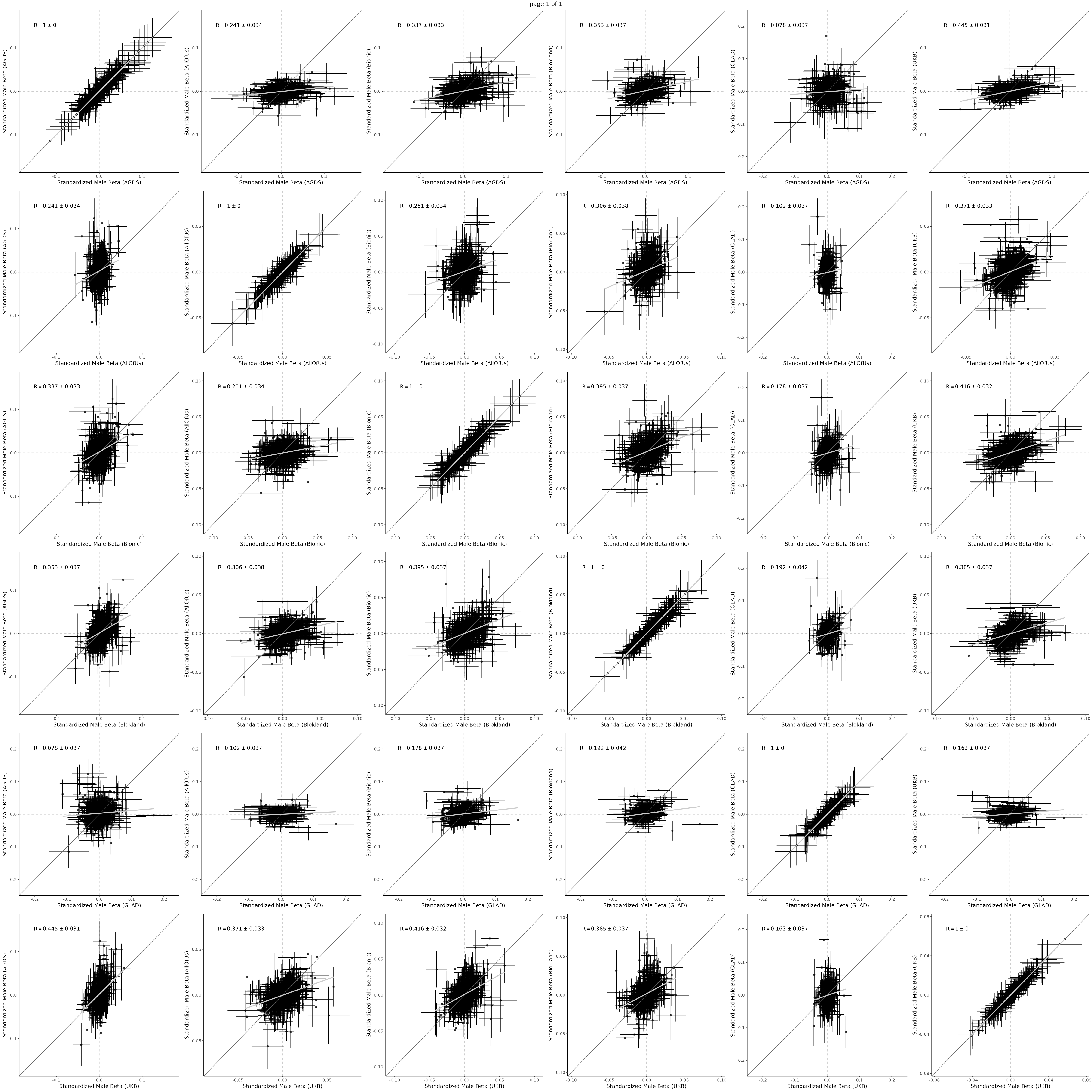

### Supplementary Figure 6

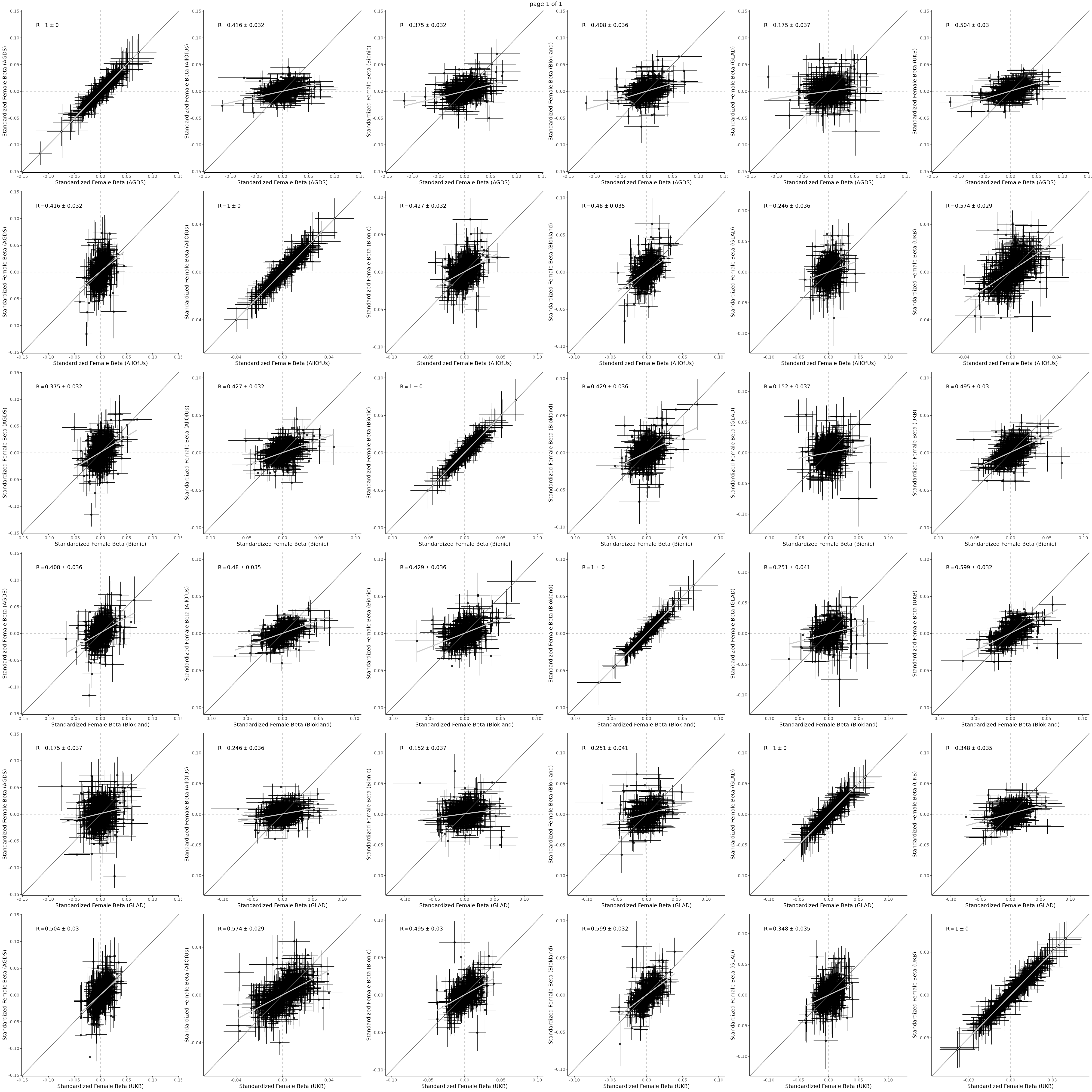

### Supplementary Figure 8

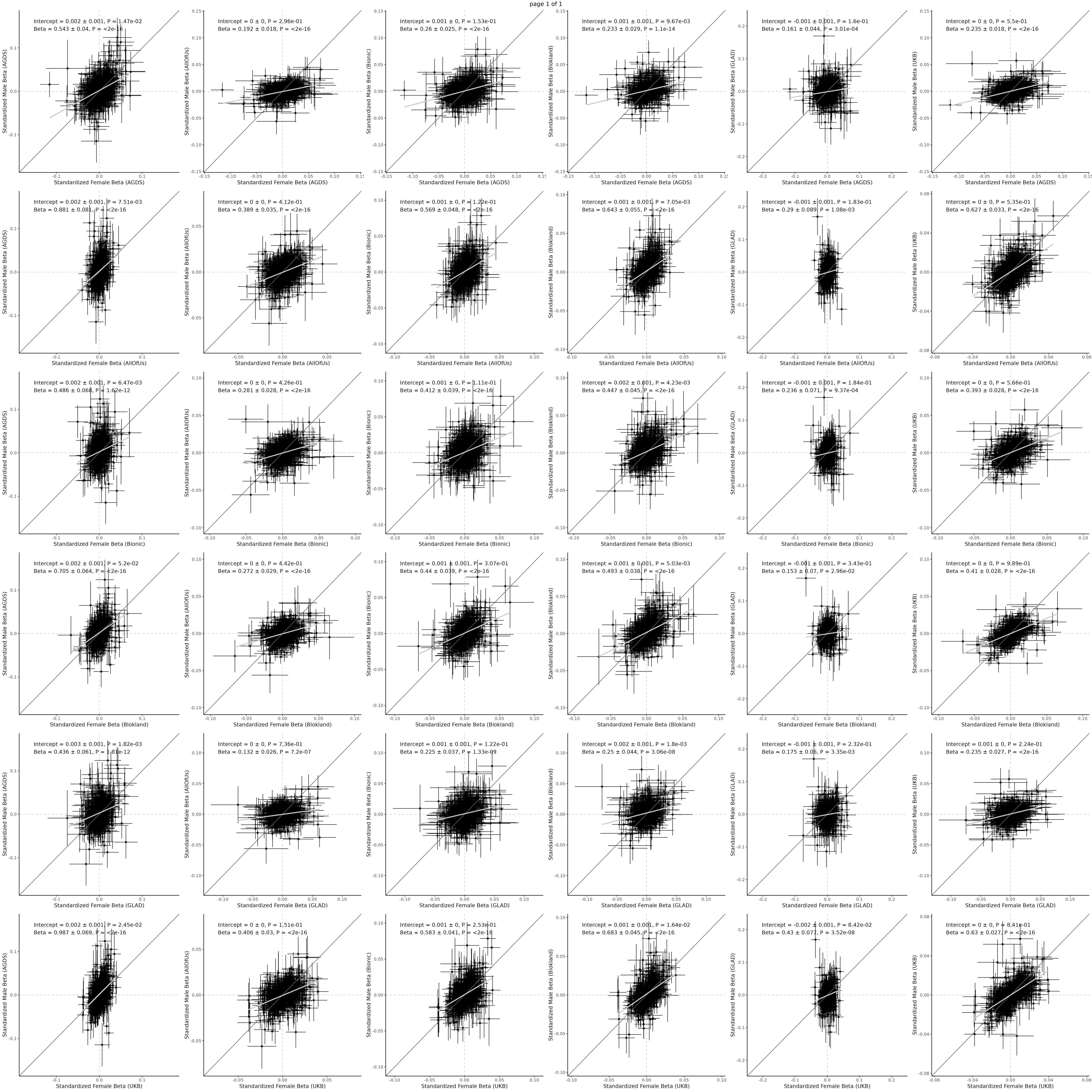

### Supplementary Figure 9

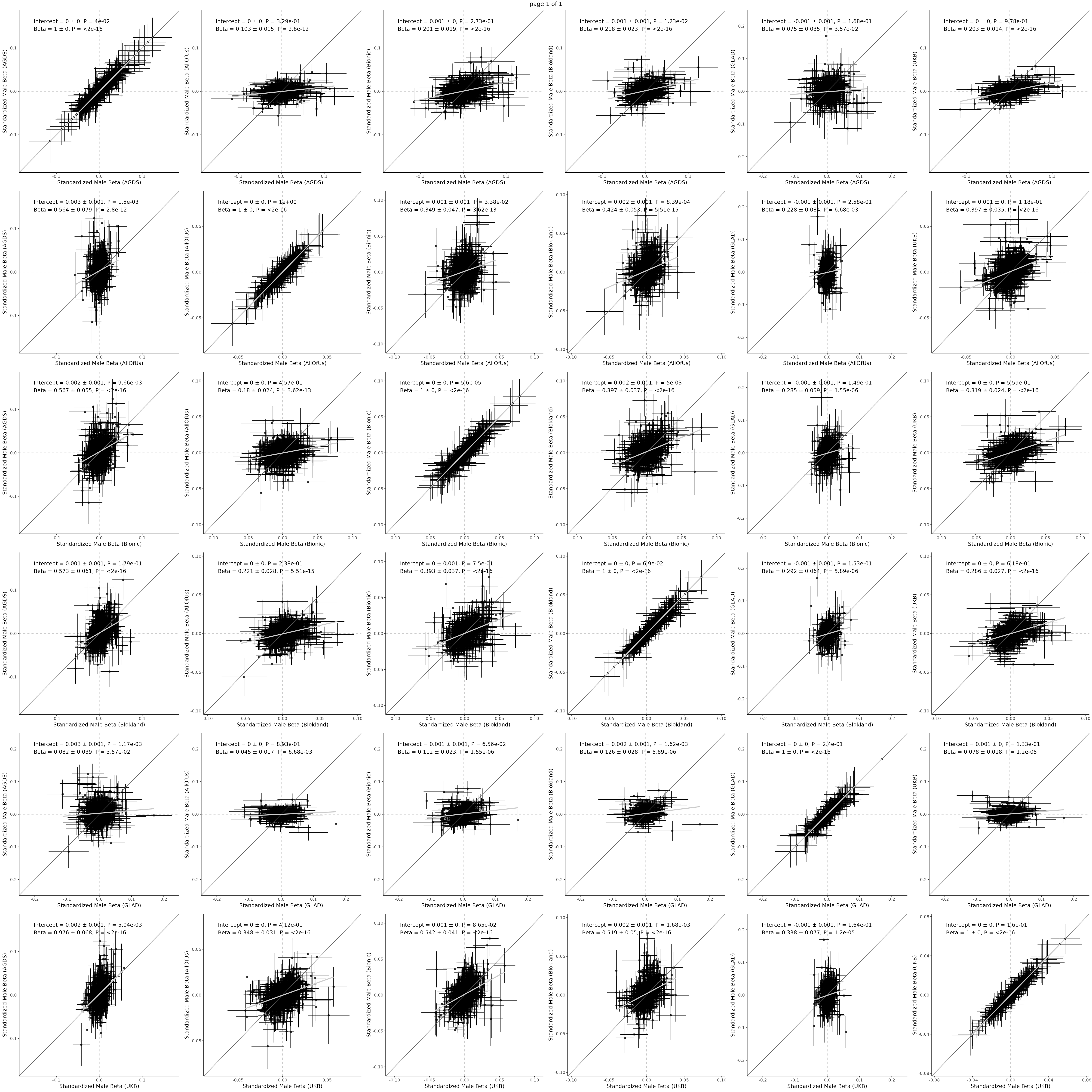

### Supplementary Figure 10

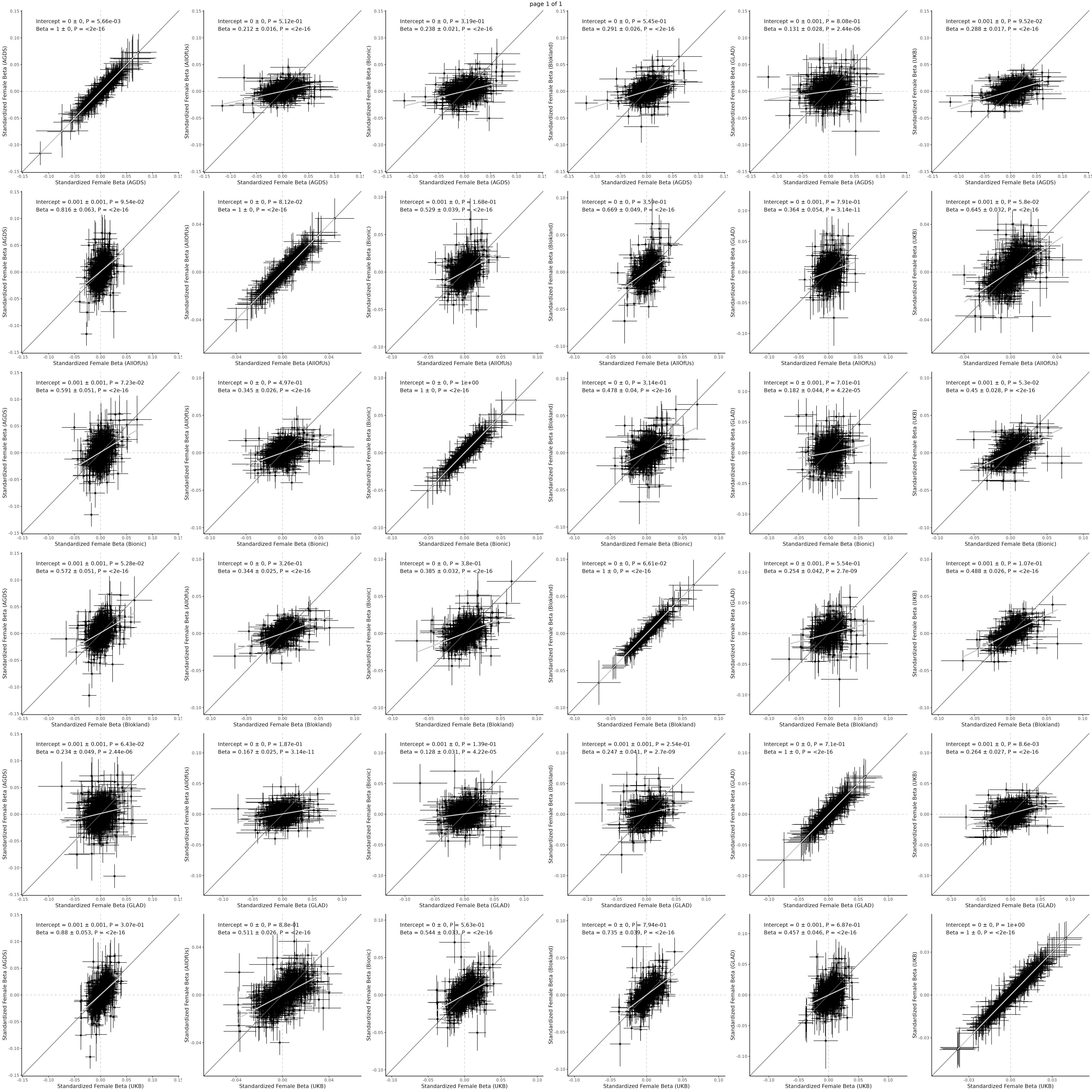
